# De novo mutation hotspots in homologous protein domains identify function-altering mutations in neurodevelopmental disorders

**DOI:** 10.1101/2022.03.19.22272594

**Authors:** Laurens Wiel, Juliet E. Hampstead, Hanka Venselaar, Lisenka E. L. M. Vissers, Han G. Brunner, Rolph Pfundt, Gerrit Vriend, Joris A. Veltman, Christian Gilissen

## Abstract

Variant interpretation remains a major challenge in medical genetics. We developed Meta-Domain HotSpot (MDHS) to identify mutational hotspots across homologous protein domains. We applied MDHS to a dataset of 45,221 *de novo* mutations (DNMs) from 31,058 patients with neurodevelopmental disorders (NDDs) and identified three significantly enriched missense DNM hotspots in the ion transport protein domain family (PF00520). The 37 unique missense DNMs that drive enrichment affect 25 genes, 19 of which were previously associated with NDDs. 3D protein structure modelling supports function-altering effects of these mutations. Hotspot genes have a unique expression pattern in tissue, and we used this pattern alongside *in silico* predictors and population constraint information to identify candidate NDD-associated genes. We also propose a lenient version of our method, which identifies 32 hotspot positions across 16 different protein domains. These positions are enriched for likely pathogenic variation in clinical databases and DNMs in other genetic disorders.

## Introduction

The interpretation of sequence variation in the context of disease remains one of the biggest challenges in genetics. *De novo* mutations (DNMs) in protein-coding genes are an established cause of neurodevelopmental disorders (NDDs)^1^, and roughly ∼45% of NDDs are caused by a DNM in a protein-coding gene.^2,3^ By modelling the probability of DNMs occurring in specific genes, one can identify genes that are enriched for DNMs in patient cohorts, provided that large enough cohorts are available. This statistical identification of NDD-associated genes requires ever-larger patient collections.^3–7^ A recent study of DNMs in 31,058 patients with NDDs concluded that NDD-association of genes still is far from saturated and that over a thousand NDD-associated genes are still to be identified.^7^

Several studies of NDD patients have found that for specific genes, missense DNMs cluster in functional regions, and that this fact can be used to identify disease-associated genes.^7–9^ Conserved protein domains are of particular interest, because they harbour ∼71%^10^ of all curated disease-causing missense variants in the Human Gene Mutation Database (HGMD)^11^ and ClinVar^12^. Indeed, missense DNMs in NDD genes are almost three times more likely located in protein domains.^7^ Clustered missense DNMs in these genes may not act through haploinsufficiency, but rather through dominant negative or gain-of-function effects.^8,9^ The detection of mutation clusters, or hotspots, can be a crucial step towards associating genes with NDDs^13^ and for gaining insight into underlying disease mechanisms.^9^

Aggregation of variation across homologous domains can be a useful method to gain insight into patterns of variation^10,14,15^ and therefore can also increase statistical power to detect hotspot positions. Methods such as mCluster^16^ and the DS-Score^17^ have been developed to detect re-occurrence of missense mutations at equivalent positions in protein domains to identify passenger mutations in cancers. In line with the same principle, we developed Meta-Domain HotSpot (MDHS), a novel method to detect mutation clustering at evolutionary equivalent positions across homologous protein domains and applied this method to DNMs from a large cohort of NDD patients to identify protein consensus positions enriched for missense variation.

## Results

### General description of the data and the processing steps

To identify hotspots of *de novo* mutations in homologous protein domains, we computed MDHS (**Equation 1**) based on unique DNMs in NDD patients (**Figure 1**). This was done for each variant type separately (missense, synonymous, nonsense). We first mapped 45,221 DNMs resulting from 31,058 patients with developmental disorders^7^ onto gene transcripts using MetaDome^14^ (**Methods**). Then, we aggregated these to 12,389 meta-domain^10^ positions (**Supplementary Data S1**). The final 15,322 DNMs represent 73.7% missense (n=11,288), 21.1% synonymous (n=3,229), and 5.3% stop-gained mutations (n=805) (**Supplementary Table 1**).

**Figure 1.**
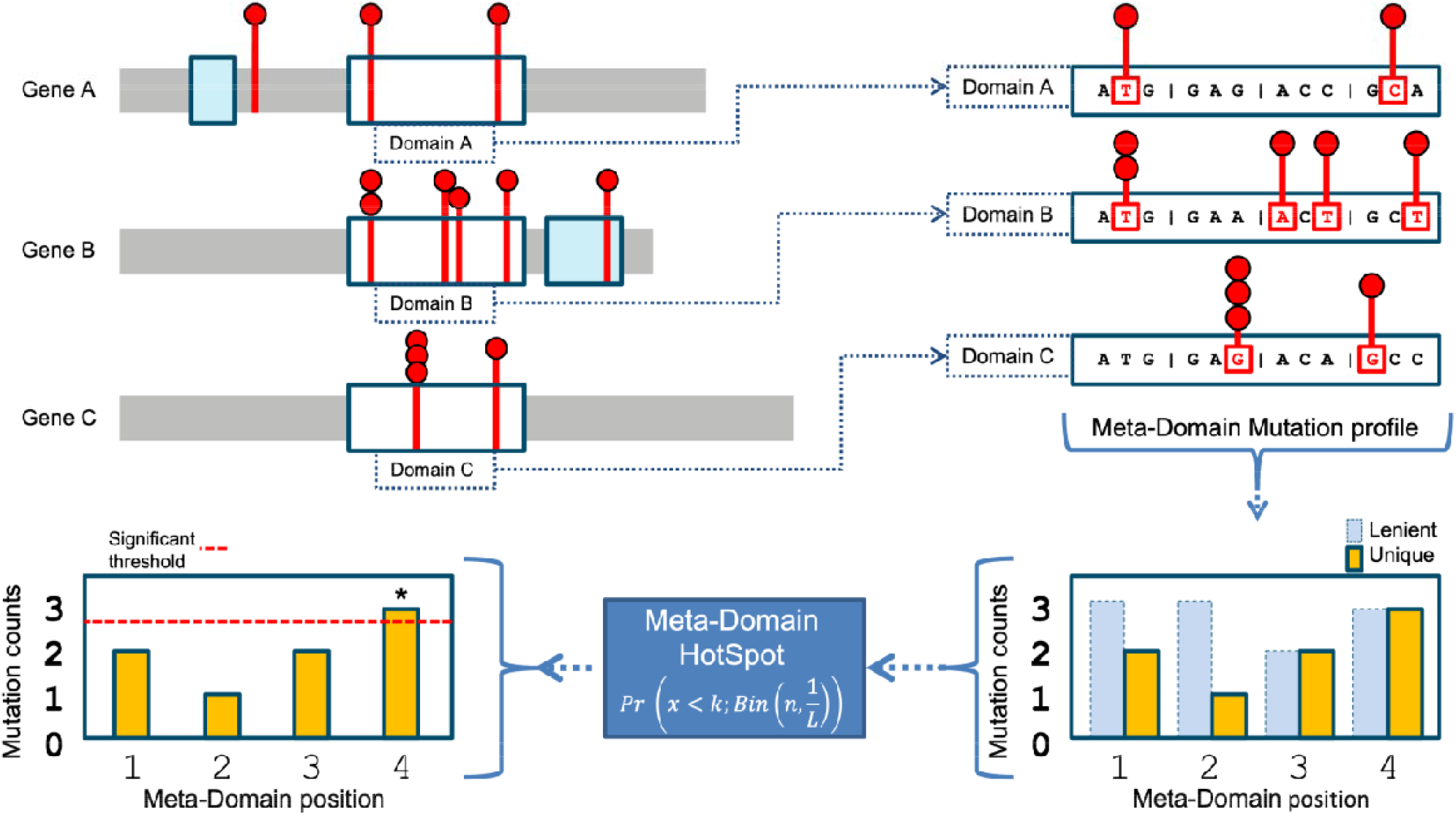
Workflow of how mutations are extracted from homologous domain regions within genes and aggregated to meta-domain positions. By clockwise orientation, starting in the upper left there are three protein representations of hypothetical genes A, B and C with the mutations identified within a cohort are displayed as red lollipops, the domains as blue and white boxes. The white boxes represent domains that are homologous and are extracted and aligned, including their mutations, and displayed on the upper right part of this image as domains A, B, and C. The mutations within a codon are then aggregated over corresponding homologous domain positions based on sequence alignments to form a meta-domain mutation profile (bottom right). Here, the recurring mutations are counted only once for unique counts (for stringent hotspot identification). The unique counts are the input for variable ‘k’ to compute a positional MDHS p-value (**Equation 1**). Together with the total number of mutations ‘n’ in the meta-domain mutation profile the significance threshold (red dotted line left bottom) can be determined which indicates a meta-domain hotspot if the mutational count exceeds it.

### Stringent DNM hotspots identified using MDHS

We initially used a stringent approach where recurrent DNMs were counted only once to prevent hotspots driven by DNMs in a single gene. Using all 11,288 missense DNMs in 2,032 protein domain families, our method identified three significant hotspots (**Supplementary Data S2-S4**) comprising 37 unique missense DNMs (57 total) in 25 different genes (**Supplementary Data S2**). Strikingly, all three hotspots are in domains belonging to the Ion Transport protein domain family (PF00520) (**Figure 2**). To validate our approach, we also performed the stringent method separately for the 3,229 synonymous and 805 stop-gained DNMs in our cohort and identified no significant hotspots (**Supplementary Data S5-S6**).

**Figure 2.**
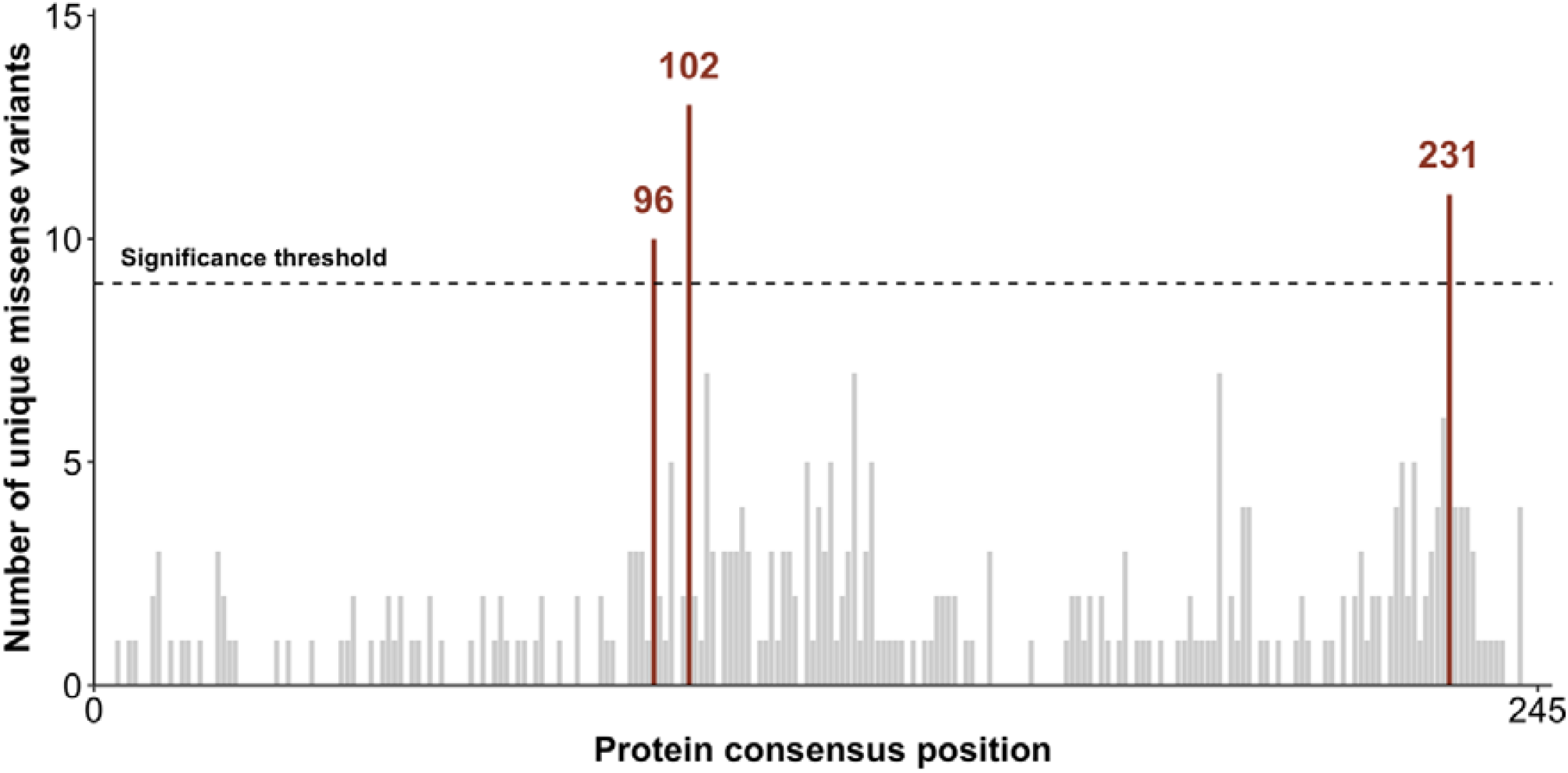
The count distribution of missense DNMs aggregated over the Ion Transport protein domain family (PF00520). The total consensus length of this domain is 245 and the sum of the count distribution is 350. The significance threshold is displayed as a dotted black line, computed via the MDHS (**Equation 1)**. The bars that exceeded the significance threshold are coloured in red and represent the mutational hotspots p.96, p.102, and p.231.

The three significant missense hotspots we identify are located on domain consensus positions p.96 (10 unique DNMs, p = 3.6 × 10^−2^), p.102 (13 unique DNMs, p = 7.1 × 10^−5^), and p.231 (14 unique DNMs, p = 7.5 × 10^−6^) of the ion transport domain family. The Ion Transport protein domain family is one of four protein domain families that we previously found to be significantly enriched with missense DNMs in NDD-associated genes.^7^ Of the 25 ‘hotspot genes’ identified with a missense DNM at a hotspot, 19 are known NDD-associated genes, representing a 3.17-fold enrichment of known NDD-associated genes (p = 1.11 × 10^−13^ chi-square test, **Supplementary Table 3**). The remaining 6 genes have not yet been associated with NDDs (**Table 1**).

**Table 1.**
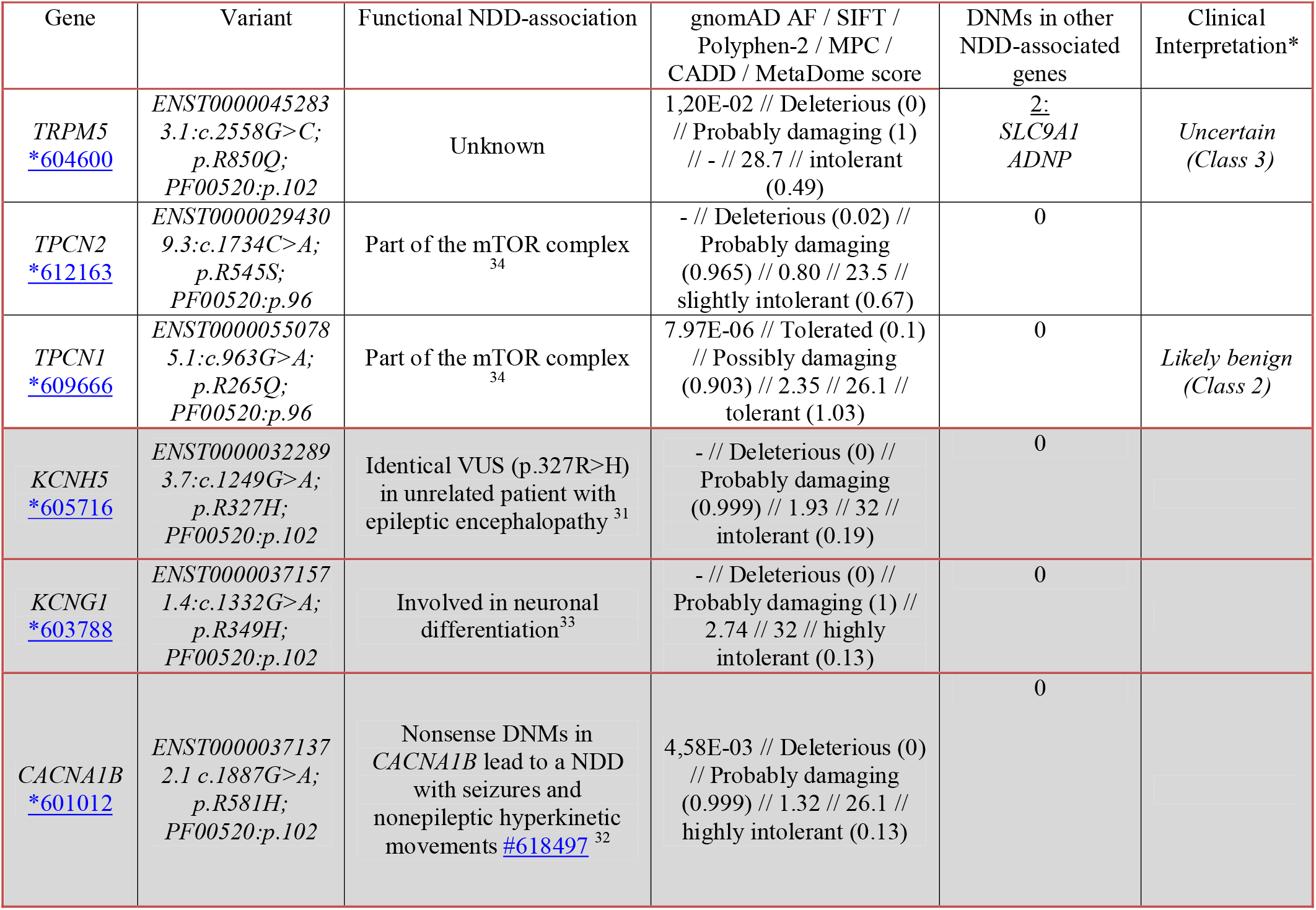
Overview of the novel genes for which we found variants at hotspot positions. First column indicates the gene and OMIM identifier. Second column indicates the identified DNM at the hotspot position. Third column indicates a previously described functional association of the gene or variant to NDD. Fourth column indicate different prediction scores of variant pathogenicity, the fifth indicates if the same patient has any DNMs located in known NDD-associated genes, and sixth ACMG classification. Rows marked in grey are novel candidate NDD genes based on additional evidence as described in this paper. Evidence for ACMG classification is provided in **Supplementary Table 6**. Genomic positions and additional phenotype information for all variants where available can be found in **Supplementary Data S3** and **S14**. **\***Variants were clinically interpreted where proband phenotype information was available (see **Supplementary Data S14**).

### Effects of missense mutations at stringent hotspots on protein structure

Mutations that cluster in genes have previously been associated with likely function-altering effects.^9^ We find that 6 out of 16 hotspot genes present in the Developmental Disorder Genotype2Phenotype (DDG2P) gene list are known to have an activating or gain-of-function mutation consequence (p = 0.0008, Fisher’s exact test), underscoring that missense mutations at hotspot positions are likely function-altering (**Supplementary Table 4**). To further investigate this, we created 3D protein structure homology models for each of the 25 genes (**Supplementary Data S1, Methods**). Ion transport protein domain 3D structures are 3-fold more identical to each other in conformation than their protein sequences would suggest (CATH-Gene3D ID: 1.20.120.350).^18^ This structural overlap encouraged us to investigate whether molecular effects of missense variants at these hotspots are likely to have similar impact on domain function across the 25 genes (**Supplementary Data S8**).

In the 25 homology models we found that hotspot p.96 (**Figure 3A**) and p.102 (**Figure 3B**) are part of the voltage-sensing helix that is important for the channel (in-)activation.^19^ These results are in line with functional studies that have been performed for missense mutations at two of these hotspots.^20,21^ Hotspot p.231 (**Figure 3C**) is part of the channel gate at the end of a transmembrane helix (**Supplementary Data S8**). In addition, we found that missense mutations follow a specific pattern for each of these hotspots. Of the 13/16 missense DNMs located at hotspot p.96 and 20/20 at p.102 change the positively charged wild-type residue to lose the positive charge. Losing positive charges at these locations has previously been described to trigger a function altering disease-mechanism (**Figure 3AB**).^20,21^ At hotspot p.231 20/21 of the missense DNMs changes the wild-type residue from a small into a larger residue. This change in residue size likely impacts the pore closure. This hypothesis is shared by Kortüm *et al*. who suggest this likely causes a steric hindrance and result into a function-altering mechanism of disease (**Figure 3C**).^22^ Overall, this shows that missense mutations at the identified hotspots are likely deleterious to domain function.

**Figure 3.**
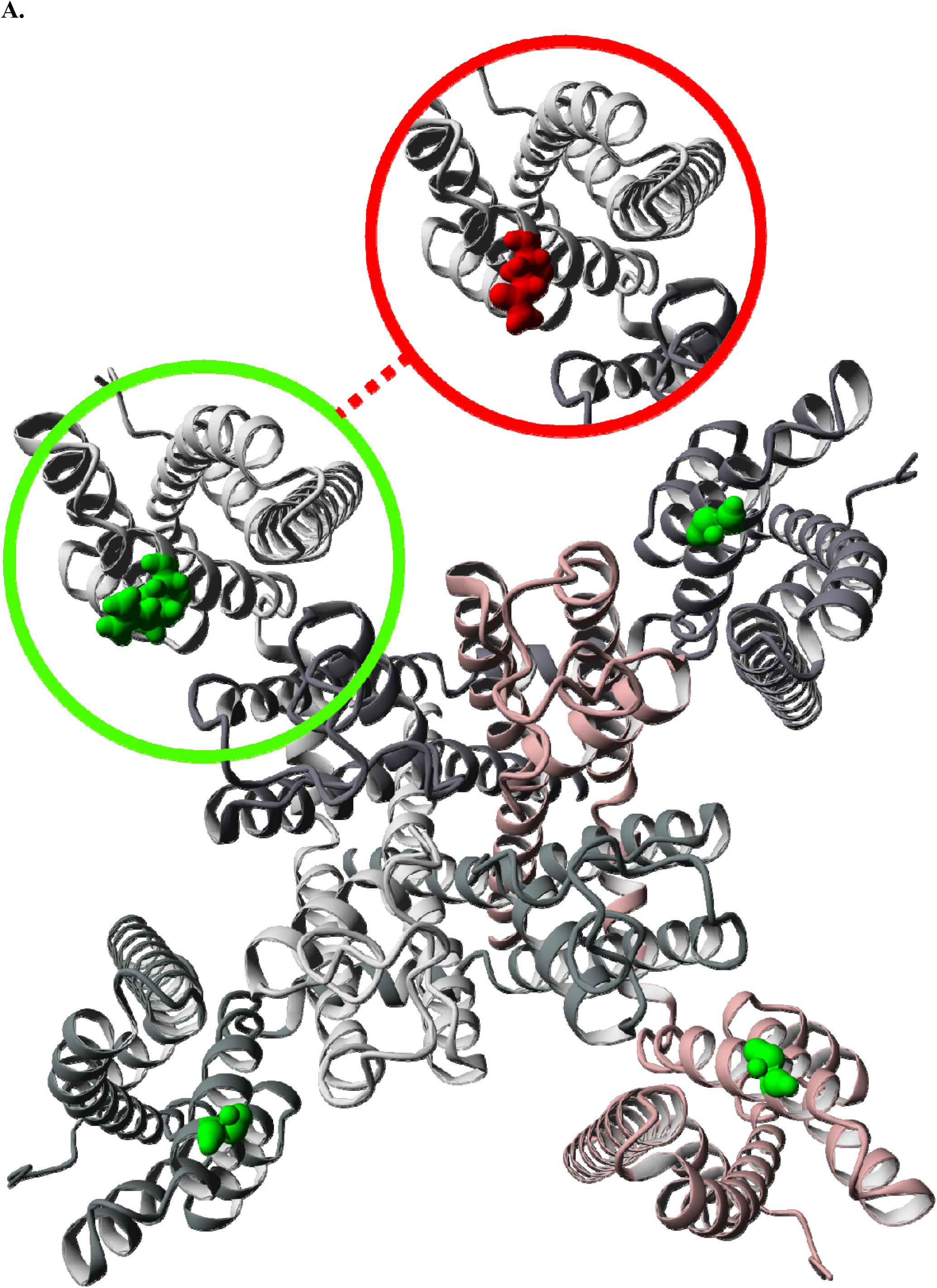
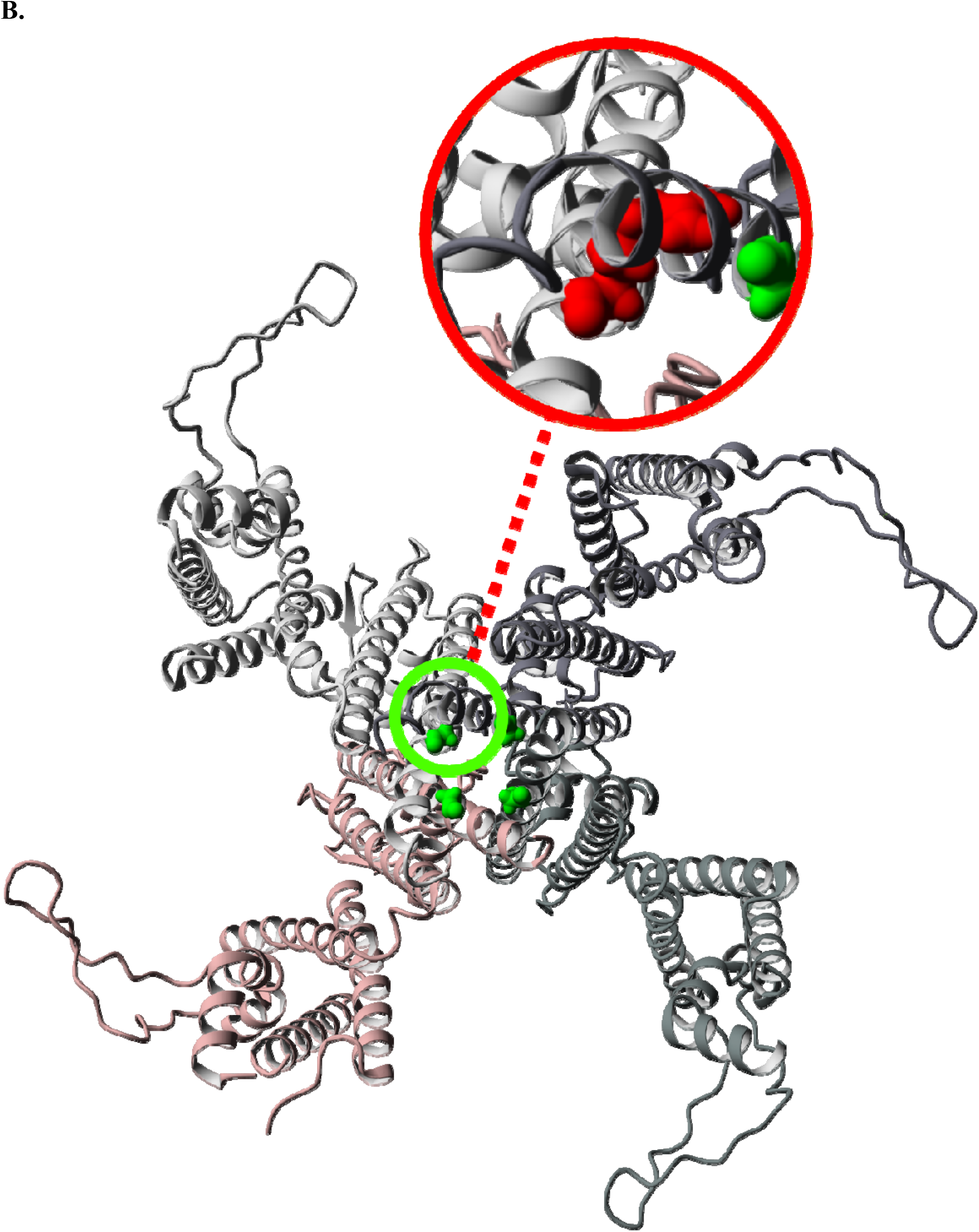
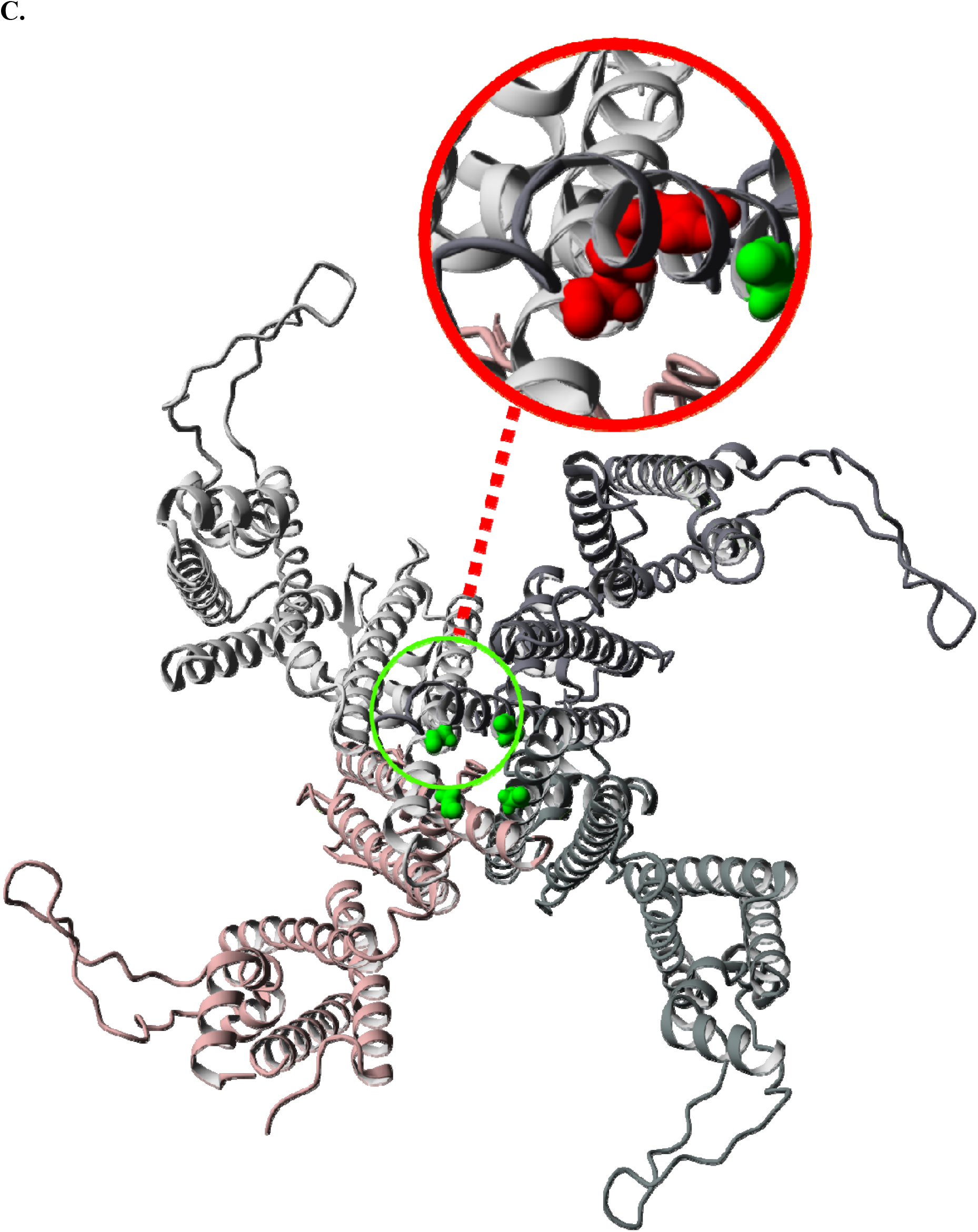
Changes in structure caused by missense DNMs in NDD-associated genes for each hotspot. **A**. Homology model of the KCNQ3 complex with missense DNM p.R227Q marked as a green to red change. The KCNQ3 complex is a tetramer constructed from four copies of the KCNQ3 monomer. All monomers are marked in different colour shades. This DNM is located at identified hotspot p.96. The wild-type Arginine residue is part of the voltage-sensing helix and changed into a Glutamine. This change causes it to lose the positive charged that was previously found to cause a function-altering mechanism of disease.^20^ **B**. Homology model of CACNA1A with missense DNM p.R1663Q marked as a green to red change. This DNM is located at identified hotspot p.102. The wild-type Arginine residue is part of the voltage-sensing helix and changed into a Glutamine. This change causes it to lose the positive charged that was previously found to cause a function-altering mechanism of disease.^21^ **C**. Homology model of the KCNH1 complex with missense DNM p.G496R marked as a green to red change. The KCNH1 complex is a tetramer constructed from four copies of the KCNH1 monomer. All monomers are marked in different colour shades. This DNM is located at identified hotspot p.231. The wild-type Glycine residue is near the pore-closing region and changed into a much larger Arginine. This may impact pore closure and is previously reported to result into a function altering mechanism of disease.^22^

### Stringent hotspot genes are constrained against missense and loss of function variation

Dominant NDD genes are characterized by population constraint against damaging genetic variation.^23,24^ We compared observed counts of loss of function, missense, and synonymous variants in control, NDD-associated, hotspot and proposed novel hotspot genes in gnomAD v2 to expected counts based on a null mutational model.^25^ Both hotspot and novel hotspot genes are constrained against loss of function and missense variation (**Figure 4A)**. Novel hotspot genes have lower constraint against loss of function variation than hotspot genes (**Supplementary Data S9)**. We also considered whether hotspots were found in regions of particular constraint against missense variation within genes. In total, 2,700 genes have statistical evidence of regional differences in missense constraint.^25^ Of these, 16 are hotspot genes, representing a significant enrichment compared to control genes (Fisher’s exact test p < 2.2 × 10^−16^) and NDD-associated genes (Fisher’s exact test p = 0.002, **Supplementary Figure 1**). Three are proposed novel hotspot genes (*KCNH5, CACNA1B, TPCN1*). Using regional missense constraint information, we show that PF00520 domains in hotspot genes are significantly more constrained against missense variation than PF00520 domains in control genes (p = 1.4 × 10^−7^, Wilcoxon rank-sum test; **Figure 4B**), but similarly constrained compared to NDD-associated genes without a hotspot that also contain a PF00520 domain (p = 0.65, Wilcoxon rank-sum test).

**Figure 4.**
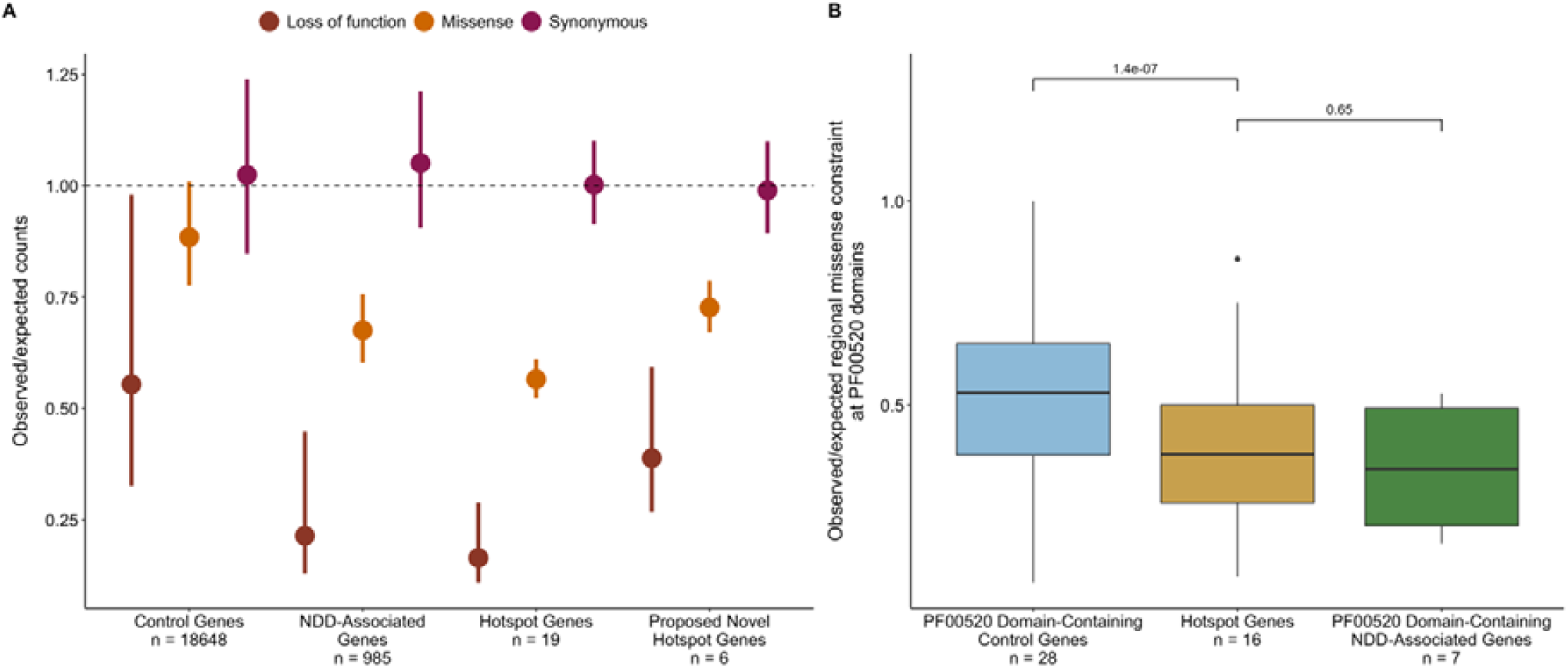
Hotspot genes are constrained against loss of function and missense variation. ***A)*** *Constraint in hotspot and proposed novel hotspot genes*. The observed variant counts for loss of function (red), missense (orange), and synonymous (pink) variants from the gnomAD v2 release were compared to the expected counts based on a null mutational model (Samocha *et. al*., 2017). Points represent the mean observed/expected ratios for all genes in each set and bars denote the mean upper and lower bound fractions for these ratios. The dashed line at observed/expected = 1 indicates perfect adherence to the null mutational model (observed counts = expected counts); values that fall below this line are constrained. ***B)*** *Mutation hotspots occur in missense constrained regions within genes*. Regional missense constraint was compared across PF00520 domain-containing control genes (blue), PF00520 domain-containing NDD-associated genes (green) and hotspot genes (yellow).

### Brain-specific expression of stringent hotspot genes

We analysed the expression of the 19 known hotspot genes in approximately 948 donors across 54 tissues from the GTEx v8 release.^26^ We observed that NDD genes and control genes have distinct gene expression patterns, with a higher proportion of NDD genes constitutively expressed across all tissues (p < 2.2 × 10^−16^, Fisher’s exact test; **Supplementary Table 5**). Hotspot genes share a characteristic expression pattern compared to these two groups (**Figure 5A**), with a significantly higher proportion of hotspot genes expressed in the brain compared to control genes and significantly lower proportion expressed in all other tissues compared to NDD genes (in 40/42 non-brain tissues, **Supplementary Data S10**). Given this tissue-specific expression pattern, we grouped GTEx tissues into two tissue groups (brain and other tissues, **Methods**). The hotspot gene set is significantly enriched for genes with higher expression in brain compared to control genes (89.4% versus 19.8% expressed higher in brain, p = 2.985 × 10^−5^, Fisher’s exact test) and NDD genes (89.4% versus 31.3%, p = 0.002, Fisher’s exact test) (**Supplementary Figure 2**). Only two hotspot genes do not have higher expression in brain: *SCN10A*, which is constitutively unexpressed across tissues in GTEx samples, and *CACNA1C* (median TPM in brain = 2.94, median TPM in other tissues = 4.35). We further show that this expression pattern is not characteristic of all genes containing an ion transport domain, but only the subset of these genes statistically associated to NDDs (**Supplementary Data S11, Supplementary Figure 3-4**).

**Figure 5.**
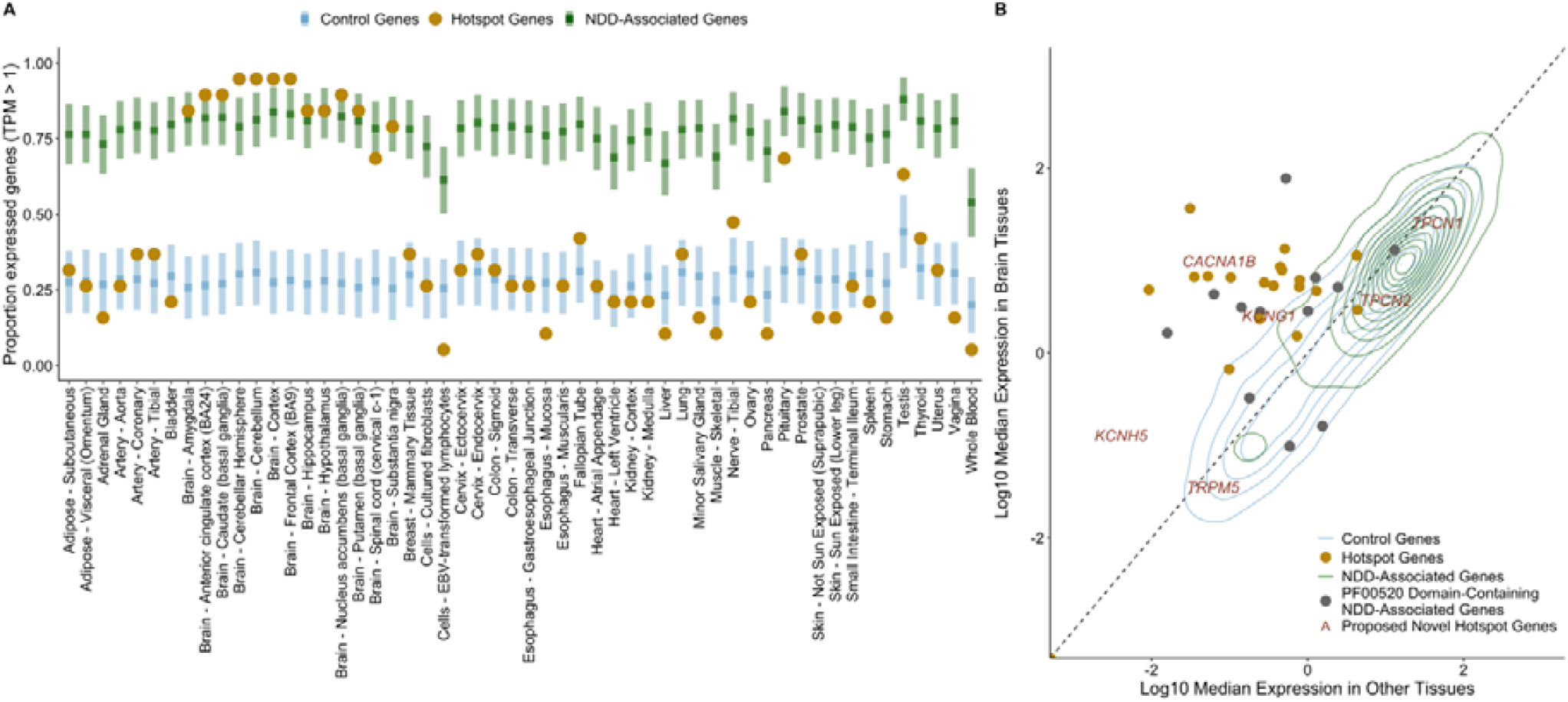
Hotspot genes have a distinct gene expression pattern. ***A)*** *Tissue expression of hotspot genes compared to control and other NDD genes*. Expression of the 19 established NDD genes containing missense DNM(s) at a stringent mutation hotspots (hotspot genes, yellow) were evaluated across 54 GTEx tissues (x-axis). Hotspot gene expression was compared to NDD genes (green) and control genes (blue). The y-axis depicts the proportion of expressed genes (**Methods**). Squares and bars depict the median and standard deviation respectively of NDD and control gene distributions. ***B)*** *TPM distribution in brain and other tissues varies across gene sets*. Control (blue) and NDD-associated (green) genes are represented by 2D density distributions. Hotspot genes (yellow) are shown as points, as are NDD-associated genes containing a PF00520 domain (grey). Proposed novel hotspot genes are marked by their gene name in red text.

We also compared the TPM distribution in brain and other tissues for control genes, NDD-associated genes, hotspot genes, and the six novel hotspot genes (**Methods; Figure 5B)**. Both hotspot and NDD-associated genes had significantly higher TPM in brain tissues than control genes (p = 0.0039 and p < 2.2 × 10^−16^, Wilcoxon rank-sum test), and both hotspot genes and control genes had significantly lower TPM in other tissues than NDD-associated genes (p < 2.2 × 10^−16^ and p = 2.08 × 10^−8^, Wilcoxon-rank sum test; **Supplementary Figure 5**). Modelling suggests that *CACNA1B* and *KCNG1* belong to the hotspot gene distribution by odds ratio (**Supplementary Data S12**). Additionally, we find 6 NDD-associated PF00520 domain-containing genes (*HCN1, TRPV3, KCNQ5, KCNC1, KCNC3, TRPM3*) that also belong to the hotspot gene distribution by odds ratio. We hypothesize that missense mutations at hotspot positions in these 6 genes may also cause NDDs.

### Lenient DNM hotspots identified using MDHS

We also implemented a lenient version of MDHS that counts all missense variants at protein consensus positions, even if they recur between individuals (see **Methods, Supplementary Figure 6**). Counting recurrent missense variants gives us more power to detect hotspot positions, but these hotspot positions may be driven by missense variation in a single domain. We detected 32 lenient missense DNM hotspots across 16 Pfam protein domain families using this method (**Supplementary Table 2**). We find a significant enrichment of genes statistically associated to NDD (Fisher’s exact *p* < 2.2 × 10^−16^) and DDG2P genes (Fisher’s exact *p* < 2.2 × 10^−16^) at lenient hotspot positions (**Supplementary Figure 7**).

We also find that missense variants at these positions are significantly more likely to be pathogenic or likely pathogenic in clinical databases (VKGL, **Figure 6A**; ClinVar, **Figure 6B**). We compared the proportion of reported likely pathogenic (LP) missense variants at hotspot positions to those at other protein consensus positions across the 16 Pfam domain families with a lenient hotspot (**Figure 6AB**). We find a significant enrichment of LP variants at hotspot positions when we consider all positions (Fisher’s exact *p* < 2.2 × 10^−16^, ClinVar; *p* < 2.2 × 10^−16^, VKGL), only positions without a DNM in our cohort (Fisher’s exact *p <* 2.2 × 10^−16^, ClinVar; *p <* 2.2 × 10^−16^, VKGL), and only codons without a DNM in our cohort (Fisher’s exact *p <* 2.2 × 10^−16^, ClinVar; *p =* 3.08 × 10^−13^, VKGL; **Supplementary Table 6**).

**Figure 6.**
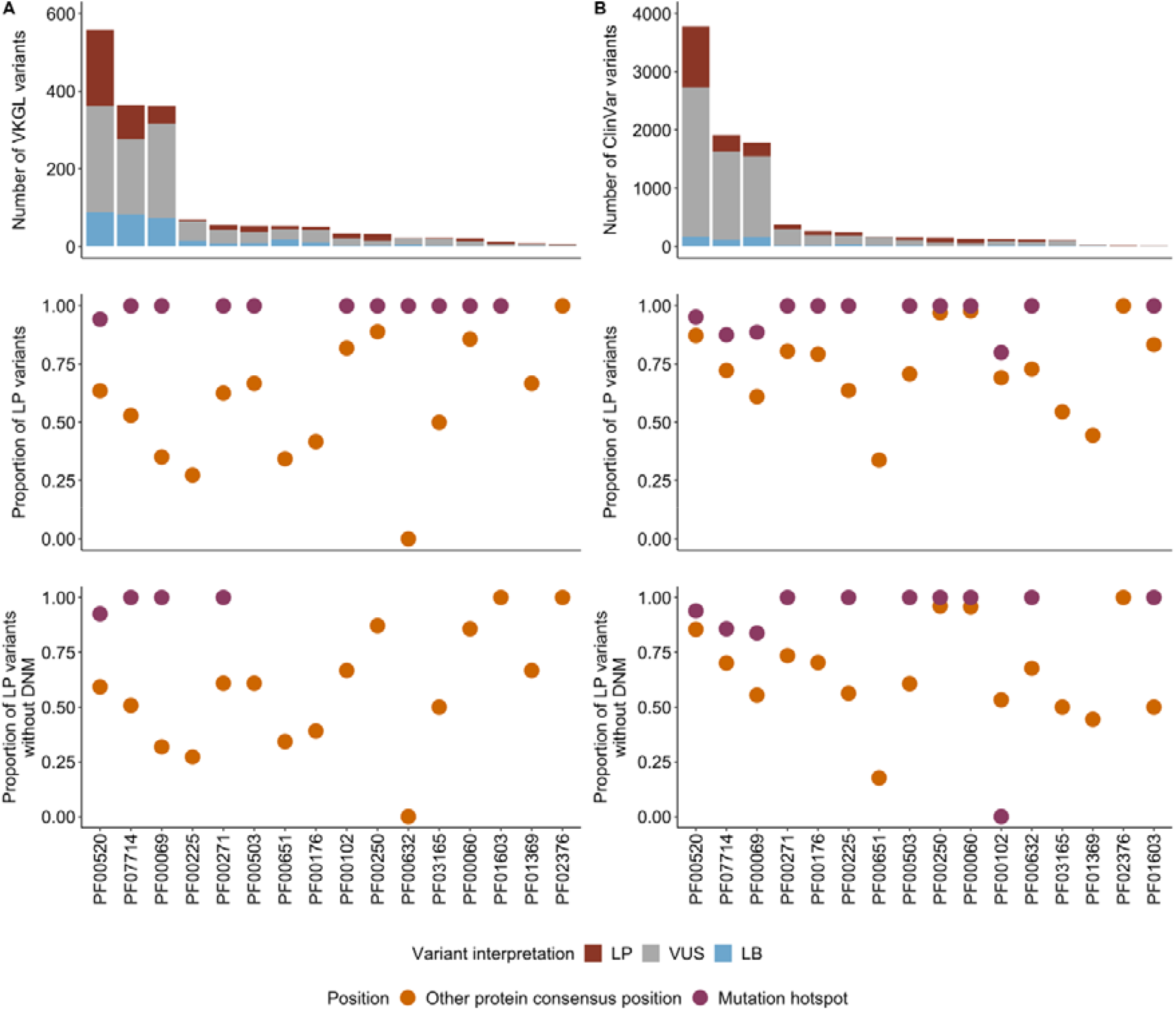
Lenient hotspots are enriched for likely pathogenic missense variation in clinical databases. Counts of likely pathogenic (LP, red), uncertain (VUS, grey) and likely benign (LB, blue) missense variants in VKGL (A) and ClinVar (B) in domains containing a lenient hotspot position. The proportion of LP missense variants (LP/(LP + LB), see **Methods**) was compared between mutation hotspots (purple) and all other protein consensus positions within the domain (orange). This comparison was done for all possible missense variants in these domains (row 2) and with positions containing DNMs in our cohort excluded (row 3).

### Lenient hotspots are enriched for missense variation in autism-spectrum disorders

We investigated if we could find further evidence for the identified lenient hotspots in a combined cohort of publicly available *de novo* mutation datasets for autism-spectrum disorders (ASD; 11,986 ASD probands, 35,584 individuals) and congenital heart defects (CHD; 2,654 trios) alongside DNMs from unaffected individuals (1,740 ASD siblings, 1,548 population controls; see **Methods**).

We observe a significant enrichment of missense DNMs at hotspot positions in NDD and ASD cohorts compared to unaffected individuals (Fisher’s exact *p* = 3.5 × 10^−13^, NDD; Fisher’s exact *p* = 0.007, ASD; Fisher’s exact *p* = 0.07, CHD; **Figure 7A, Supplementary Table 7**). We observe no significant enrichment in synonymous DNMs at hotspot positions in any cohort (**Supplementary Table 8**). However, we predict that some lenient hotspot positions are driven by mutational processes or ascertainment bias in particular genes and not necessarily by the cumulative effect of pathogenic mutations across several genes. To correct for this, we also tested for an enrichment of missense variants unique to ASD and CHD probands at lenient hotspots (**Figure 7B, Supplementary Table 9**). We found a significant enrichment of these unique missense variants in ASD probands (Fisher’s exact *p* = 0.047) but not in CHD probands (Fisher’s exact *p =* 1). The majority (10/13, 77%) of the missense variants driving this enrichment in ASD probands are in genes statistically associated to NDDs.^7^

**Figure 7.**
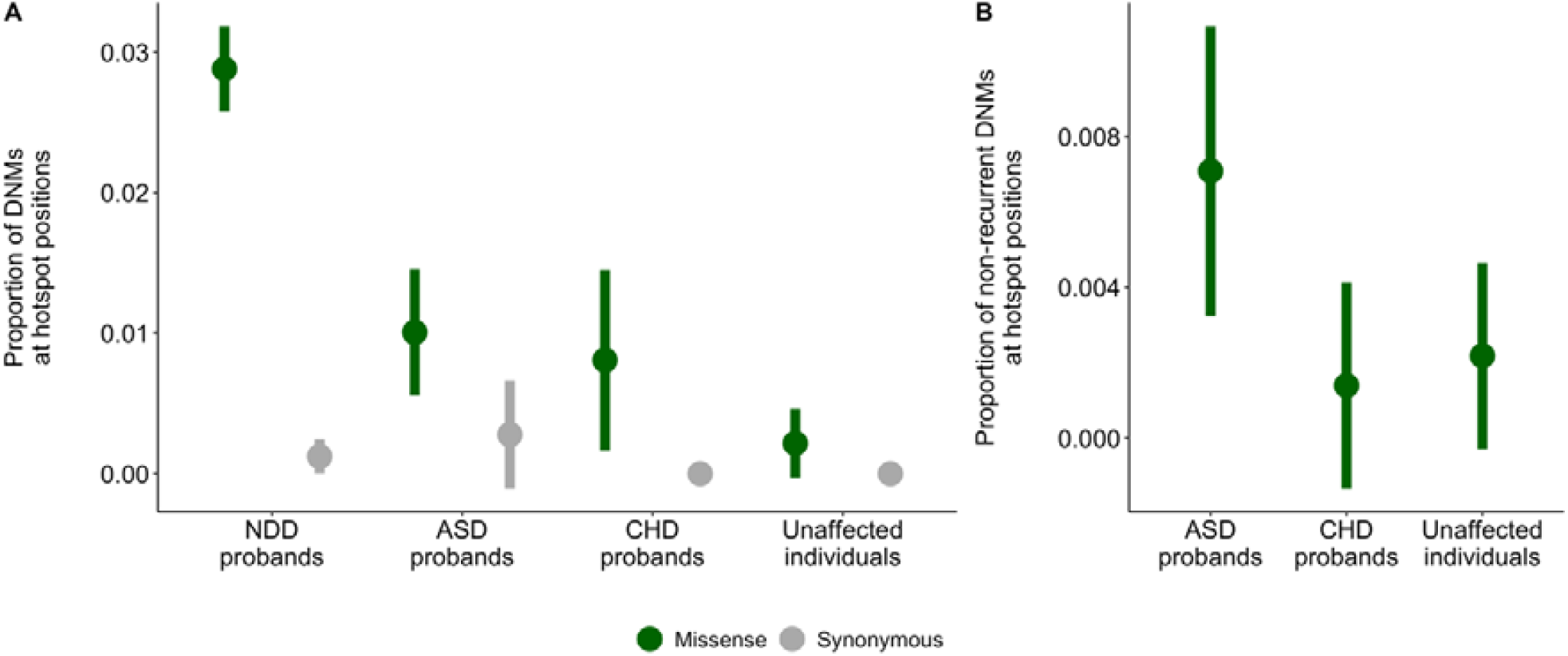
Patients are enriched for missense variants in lenient hotspot positions compared to healthy population controls. A) NDD and ASD are enriched for missense DNMs in lenient hotspot positions compared to unaffected individuals (green) but are not enriched for synonymous DNMs (grey). Only DNMs within protein consensus positions were used for this comparison (see **Methods**). B) ASD probands are enriched for missense DNMs in lenient hotspot positions (green) not present in our NDD cohort.

## Discussion

By exploiting homology within the human genome we were able to identifyied mutational clustering of DNMs at evolutionarily conserved positions across genes that share protein domains. We identify identified three stringent (p.96, p.102, p.231) and 32 lenient mutational hotspots across 16 Pfam domain families using our MDHS method. Missense DNMs at stringent hotspots are located in 25 genes within our cohort. Structural and functional work by us and others suggest that missense mutations at these positions may be gain-of-function.^27,28^

The hotspots we statistically identify in our cohort may have broader clinical relevance. We hypothesize that the 19 hotspot genes are examples of a broader class of ion transport domain-containing genes, and that missense mutations at hotspot positions in these genes are generally damaging. Our finding that clinical databases contain many pathogenic missense mutations at hotspot positions in other monogenic disease genes (**Supplementary Data S13**) support this idea. Other studies have shown that missense mutations in ion transport domain-containing genes may have position-specific functional effects.^29,30^ Several of these mutations occur in genes not statistically associated to NDDs, indicating that missense mutations at hotspot positions could be pathogenic across a variety of disorders. In line with this, we observe that some PF00520 domain-containing NDD-associated genes have lower expression in brain but have a similar level of tissue-specific expression in a non-brain tissue. *SCN4A*, for example, is not expressed in brain and is predominantly expressed in skeletal muscle. Although the expression pattern of *SCN4A* is different from the hotspot genes presented in our analysis, hotspot positions in *SCN4A* are similarly constrained against missense variation (**Figure 5B**). We hypothesise that phenotypes associated with pathogenic mutations at hotspot positions may vary depending on where the mutated gene is expressed. For example, of the four PF00520 domain-containing genes predominantly expressed in skeletal muscle (*SCN4A, CACNA1S, RYR1*, and *KCNA7*), three of these (*SCN4A, RYR1*, and *CACNA1S*) have pathogenic missense variation at hotspot positions in clinical databases (**Supplementary Data S13**). Patients with these mutations present with disorders of the skeletal muscle, including myotonia, paramyotonia and hyperkalemic paralysis. Our work suggests that missense mutations at hotspot positions in *KCNA7* may also result in skeletal muscle disorders based on the tissue expression of *KCNA7* and the conservation of hotspot positions in the PF00520 domain of this gene.

Our method also identified six genes with a mutation at the hotspot location that have not previously been associated with NDDs. Three genes – *KCNH5, CACNA1B*, and *KCNG1* – have evidence supporting NDD association (**Supplementary Table 10**). In *KCNH5*, the same DNM was described as a variant of unknown significance (VUS) in a patient with an epileptic encephalopathy,^31^ and studies are ongoing that associate this gene with NDD in other patients (personal communication Prof. Heather Mefford). *CACNA1B* was recently established as a NDD-associated gene on the basis of LoF DNM enrichment.^32^ In line with this, *CACNA1B* is the only proposed novel hotspot gene that is predicted to be intolerant to heterozygous loss of function by population constraint (pLI = 1; **Supplementary Data S9**). However, our work suggests that the missense variants we identify at hotspot positions in *CACNA1B* may be function-altering. *KCNG1* has been implicated in neuronal development^33^, and the expression profile matches well with that of the other NDD genes that have hotspot mutations. There is also some circumstantial evidence for two of the other three genes. Both *TPCN1* and *TPCN2* have no prior NDD-association, however, both genes are part of the mTOR complex, which has previously been associated to NDDs.^34^ Phenotypic data for the patient with the missense DNM in *TPCN1* shows that this patient has macrocephaly and severe ASD (**Supplementary Data S14**) which is in line with the fact that mTOR genes have been associated with intracranial volume and intellectual disability.^35^ However, the only *TPCN1* missense mutation described in literature at a hotspot position (p.102) is associated with early-onset cardiomyopathy.^36^

In this analysis, we initially identified stringent mutation hotspots statistically using unique missense mutation counts. While MDHS analysis on even larger cohorts may identify additional hotspots, we believe our method can be used on smaller datasets by also considering recurrent mutations. Applying this lenient method to our cohort identified 32 significant missense hotspots (**Supplementary Data S3)** and no significant hotspots for synonymous or nonsense mutations (**Supplementary Data S4-S5**). 12 protein domain families had hotspots spanning multiple gene-codons based on 245 DNMs from 67 genes. 48 of these 67 genes (72%) are statistically associated with NDDs, representing a 2.53-fold enrichment (p = 1.26^-31^ chi-square test; **Supplementary Table 11)** and showing the merit of this approach. While the inclusion of recurrent mutations allows hotspots driven by proliferative advantages of single mutations in the germline or soma, CpG hypermutability, or biases in clinical ascertainment, it also increases power to detect robust hotspot positions (**Supplementary Figure 6**). In support of this, we find an enrichment of likely pathogenic missense variants at lenient hotspot positions in clinical databases even if we look only at codons without a mutation in our cohort, showing the merit of this approach. However, additional filtering may be required to remove hotspots driven solely by recurrent missense mutations. Additionally, there is in principle no reason to restrict this method to *de novo* mutations; it could easily be applied to rare inherited variants in large patient cohorts.

Kaplanis *et al*. estimate that approximately 350,000 parent-offspring trios would be required to detect the majority of remaining haploinsufficient genes associated with NDDs. Genes with gain-of-function effects are predicted to be even more difficult to detect, even in very large cohorts. Methods based on homology like MDHS provide a novel approach to the identification of disease genes and mechanisms in existing datasets without increasing cohort size.

## Materials and Methods

### Dataset of *de novo* mutations

We obtained the set of 45,221 DNMs from the Kaplanis *et al* study.^7^ These DNMs were identified in 31,058 DD patients combined across three centres. The genetic testing approach of these patients were described previously per centre: DDD,^3^ GeneDX,^37^ and, Radboudumc.^4^ All individuals that underwent genetic testing provided informed consent.^7^ Subsets of these patients have been analysed and reported in previous publications.^3,23,37,38^

### Developmental disorder diagnostic gene lists

We use the diagnostic lists of DD-associated genes from the Kaplanis *et al*. study. We consider all genes statistically associated to NDDs in this study to be NDD genes (novel, consensus, and discordant genes, total genes = 1010).^7^

Additionally, we used the Deciphering Developmental Disorders Genotype2Phenotype (DDG2P, accessed 22-04-2021) list to assess the burden of gain of function mutational mechanisms already described in hotspot gene families. We considered activating, gain of function, dominant negative, and increased gene dosage mutation consequences in this list to be gain of function.

### Annotation of transcript details, protein and meta-domain position annotation

The DNMs were annotated with corresponding GENCODE^39^ transcripts from release 19 GRCh37.p13 Basic set, protein information from UniProtKB/Swiss-Prot^40^ Release 2016_09, Pfam-A^41^ v30.0 protein domains information, and meta-domain^10^ positions using a local version of the MetaDome^14^ web server (code available at https://github.com/cmbi/metadome). Meta-domains are multiple sequence alignments of regions within human protein-coding genes that correspond to Pfam protein domain families. The DNMs that correspond to Pfam consensus positions are annotated with the corresponding Pfam domain ID and consensus position.

### Filtering the annotated DNMs

The annotation process can result in multiple GENCODE gene transcripts per DNM. To ensure a single GENCODE transcript per gene we performed a filtering step by the following order of criteria:

1. Filter to variants that with transcript consequence: missense, synonymous, or, stop-gained
2. The transcript corresponds to a human canonical or isoform entry in Swiss-Prot
3. This transcript contains all (or most) of the *de novo* mutations for the corresponding gene
4. The transcript translates to the longest protein sequence length
5. If multiple transcripts remain for a gene, one of these is selected
6. Filter variants only to those that are in a Pfam protein domain

### MDHS: Detection of variant hotspots in homologous protein domains

The Pfam domain ID and consensus position allows for aggregation of genetic variants through meta-domain positions. To identify meta-domain positions that are significantly enriched with variants we created the MDHS (Meta-Domain HotSpot) p-value as follows:

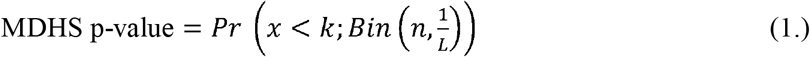

In the context of meta-domains, *n* corresponds to the total number of aggregated genetic variants for the Pfam domain ID, *L* is the total number of possible consensus positions for a Pfam domain ID, *k* is the total number of genetic variants aggregated at a single consensus position, and *x* = *k* − 1, which depicts the chance of finding less than observed genetic variants at the consensus position. The MDHS p-value is adapted from the mCluster^16^ and DS-Score^17^. In line with these methods, variants are assumed to follow a Binomial distribution. We correct the MDHS p-value via the Bonferroni method for the total number of Pfam protein domain IDs considered. If a Bonferroni corrected MDHS p-value < 0.05, we consider it as a significant mutational hotspot.

Our code to analyse the MDHS p-value was optimized to only compute for domains which can have significant hotspots. It implements a filter for domain families which works as follows: 1.) Count the number of domain consensus positions with one or more variant as ‘*n_hotspot_candidates*’. 2.) Count the number of DNMs that span at least more than one unique protein position at each consensus position and sum them up to represent ‘*n_hotspots_with_variation_from_more_then_one_protein_position*’. 3.) Apply the filter criteria such that each domain family abides:

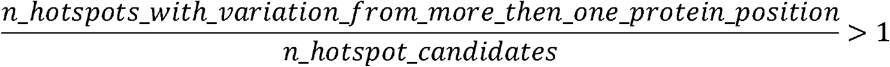

See the value at column ‘hotspot_uniqueness’ in **Supplementary Data S2, S5, S6. Stringent and lenient counting of variants in MDHS**

We use two ways to determine variable *k* in the MDHS p-value (**Equation 1**). Unless otherwise specified, we count unique variants by considering mutated chromosomal positions only once, thereby reducing the impact of recurrent mutations in a single gene (stringent). Alternatively, we refer to a lenient way of variant counting when we count every mutation equally (including recurrent mutations). For a schematic, see **Figure 1**.

### Functional characterization

We used the Ensembl Variant Effect Predictor (VEP)^42^ to annotate gnomAD allele frequency (AF)^24^, SIFT^43^, Polyphen-2^44^, MPC^25^, and the CADD_Phred^45^. MetaDome^14^ tolerance indication based on regional d_N_/d_S_ was obtained manually. ACMG^46^ classification was obtained through variant curation by a laboratory specialist. Available phenotype information for patients with missense mutations in hotspot positions can be found in **Supplementary Data S14**.

### Protein 3D structure modelling of the genes with identified hotspots

For each of the 25 genes with a DNM located at one of the hotspots we submitted the corresponding protein sequence (based on the transcript of **Filtering the annotated DNMs**) to the YASARA & WHAT IF Twinset^47,48^ homology modelling script using the default settings. The regions corresponding to the PF00520 were extracted from all resulting homology structures and combined in a single YASARA scene and then structurally aligned using the MOTIF script. The structures are available in **Supplementary Data S7**. The protein structure effects of mutations have been reported in **Supplementary Data S8**.

### Population constraint

Constraint information, including observed and expected counts, z-scores, and pLI, pNull, and pRec were calculated on gnomAD v2.1.1.^24^

### Regional missense constraint

Genes with regions of differential missense constraint were identified as described by Samocha *et. al*, 2017 in the ExAC^49^ dataset. In brief, the fraction of observed missense variation along a transcript was tested for uniformity using a likelihood ratio test. If the distribution was not uniform, the transcript was considered to have evidence of regional missense constraint. 2,700 genes showed evidence of at least two regions of distinct missense constraint using this method.

### Expression data

Pre-computed tissue expression values in transcripts per million (TPM) were taken from GTEx v8 (GTEx_Analysis_2017-06-05_v8_RNASeQCv1.1.9_gene_median_tpm.gct.gz).^26,50^

### Gene sets for constraint and expression analysis

The set of 56,200 genes for which median TPM values were available was divided into one of four sets: proposed novel hotspot genes, hotspot genes, NDD-associated genes, and control genes. Genes containing a mutation hotspot were divided into two categories: hotspot genes, n = 19, and proposed novel hotspot genes, n = 6. These categories were distinguished by presence on the DD gene list (see **Developmental disorder diagnostic gene lists**); hotspot genes are present on this list while proposed novel hotspot genes are not. The remaining genes were divided into NDD-associated genes (n = 992, excluding hotspot genes) and control genes not statistically associated with intellectual disability or developmental delay (n = 55,183). For some analyses, control genes present in DDG2P (n = 1,250, accessed 22-04-2021) or OMIM (n = 3,402, accessed 31-08-2021) but not statistically associated to NDDs were considered separate classes. Additionally, only some of these genes had population constraint information available from gnomAD v2.1.1 (n = 19,658). Genes in all sets can be found in **Supplementary Data S15**.

Of the 56,200 genes described above, 105 contained a PF00520 domain in MetaDome v1.0.1. These 105 genes represented 19 hotspot genes, 6 proposed novel hotspot genes, 12 NDD-associated genes not containing a missense DNM at a hotspot position in our cohort, and 68 control genes (of which 6 are present in DDG2P and 26 in OMIM; **Supplementary Data S16)**.

### Proportion of expressed genes across GTEx tissues

A fixed level of TPM > 1 was used to define expression in each tissue. NDD-associated and control genes were randomly sampled 1000 times into sets of 19 genes, and the proportion of expressed genes (number of genes with TPM > 1/total number of genes) was calculated for each set. This generated a distribution of proportions across 54 GTEx tissues. The proportion of expressed hotspot genes per tissue was computed without sampling.

### TPM differences between tissue groups

In order to assess expression differences between brain and other tissues, GTEx tissues were divided into two groups. We considered the amygdala, anterior cingulate cortex (BA24), caudate (basal ganglia), cerebellar hemisphere, cerebellum, cortex, frontal cortex (BA9), hippocampus, hypothalamus, nucleus accumbens (basal ganglia), putamen (basal ganglia), spinal cord (cervical c-1), and substantia nigra brain tissues (n = 12). All non-brain tissues were included in the ‘other tissues’ set (n = 42). The TPM value for each tissue set was defined as the median TPM of all tissues in the set. Based on these differences, we modelled the brain TPM distribution in hotspot genes and control genes and the other tissue TPM distribution in hotspot genes and NDD-associated genes as normal distributions. For each set of distributions, a likelihood ratio of belonging to each distribution was calculated. Proposed novel hotspot genes were considered to have evidence for association with NDDs if they were more likely to belong to the hotspot gene distribution across both tests.

### Filtering and annotation of additional *de novo* mutation cohorts

We analysed the enrichment of missense and synonymous DNMs in lenient hotspot positions across a total of three additional published DNM cohorts. We used an autism-spectrum disorder (ASD) cohort published by Satterstrom *et al*. (35,584 total individuals, 11,986 with ASD), a congenital heart defect (CHD) cohort published by Jin *et al*. (2,645 trios), and a cohort of healthy individuals sequenced by Jonsson *et al*. (1,548 trios). To increase our power to find significant differences at lenient hotspot positions, we pooled the Jonsson *et al*. healthy individuals with unaffected siblings from the Satterstrom ASD cohort (1,740 siblings) for a total of 3,288 unaffected individuals.

Annotation of meta-domain protein consensus positions for these datasets was done as previously described for Kaplanis *et al*. using MetaDome. The number of PTV, missense, and synonymous SNVs in Pfam protein domains in each dataset can be found in **Supplementary Table 12**.

### Variant annotation

Variants at protein consensus positions were checked for clinical interpretation across four curated variant databases: ClinVar, HGMDPro, Swiss-Prot and VKGL, all accessed 21-08-2021. Mapping of protein consensus positions to GRCh37 genomic positions for each gene as done using MetaDome.

For analysis on stringent hotspot positions, ClinVar data was unfiltered on evidence level or review status. We classified missense variation as LP (pathogenic or likely pathogenic, ACMG Class V or IV) or VUS (variant of uncertain significance, ACMG Class III) based on the most severe class across all four databases (Supplementary Data S13).

For the enrichment analysis on lenient hotspot positions, only ClinVar and VKGL data was used because these two databases include likely benign variants. ClinVar data was filtered on review status (required to be one of ‘practice guideline’, ‘reviewed by expert panel’, ‘criteria provided, multiple submitters, no conflicts’, ‘criteria provided, single submitter’). Variants were classified as LP (ACMG Class V or IV), VUS (ACMG Class III) or LB (benign or likely benign, ACMG Class II or I). Variants with conflicting interpretations of pathogenicity within or between databases (LP and LB annotations) were removed, as were variants with only VUS annotations.

## Data Availability

Most data produced in this study are contained in the manuscript. All data produced in this study are available upon reasonable request to the authors.

https://stuart.radboudumc.nl/metadome/

https://www.gtexportal.org/home/

## Code availability and data access

Constraint and expression analysis was done in R version 3.6.2. Code and data to reproduce all parts of the analysis is available on GitHub (https://github.com/cmbi/MetaDomainHotSpot). MetaDome was used to annotate meta-domains, the source code for this is also available on Github (https://github.com/cmbi/metadome).

## Acknowledgements

We thank Dr. Torti and Dr. Retterer from GeneDX for connecting us with Prof. Mefford. We thank Prof. Mefford for information regarding the likely NDD-association of *KCHN5*. We thank Elke de Boer for useful discussions. This work was in part financially supported by grants from the Netherlands Organization for Scientific Research (916-14-043 to C.G. and 918-15-667 to J.A.V.), and from the Radboud Institute for Molecular Life Sciences, Radboud University Medical Center (R0002793 to G.V.).

## URLs

YASARA: http://www.yasara.org/

CATH-Gene3D: http://www.cathdb.info/

Variant Effect Predictor: https://www.ensembl.org/Tools/VEP

GTEx: https://www.gtexportal.org/home/

MetaDome web server: https://stuart.radboudumc.nl/metadome/

MetaDome GitHub repository: https://github.com/cmbi/metadome

MetaDomainHotspot analyses repository: https://github.com/cmbi/MetaDomainHotSpot

DECIPHER: https://www.deciphergenomics.org/

HGMD: http://www.hgmd.cf.ac.uk/ac/index.php

ClinVar: https://www.ncbi.nlm.nih.gov/clinvar/

Swiss-Prot: https://www.uniprot.org/

VKGL: https://www.vkgl.nl/nl/diagnostiek/vkgl-datashare-database

## medRxiv Note

Our **Supplementary Data S7** file (YASARA protein structures) cannot be made available through medRxiv. Please contact the corresponding author for this data.

## Notes

### Competing Interest Statement

The authors have declared no competing interest.

### Funding Statement

This study was in part financially supported by grants from the Netherlands Organization for Scientific Research (916-14-043 to C.G. and 918-15-667 to J.A.V.), and from the Radboud Institute for Molecular Life Sciences, Radboud University Medical Center (R0002793 to G.V.).

### Author Declarations

This study involves only openly available human data. Developmental disorder (NDD) DNMs: Kaplanis et al., Nature, 2020. Autism-spectrum disorder (ASD) DNMs: Satterstrom et al., Cell, 2020. Congenital heart defect (CHD) DNMs: Jin et al., Nature Genetics, 2017. DNMs from unaffected individuals: Jonsson et al., Nature, 2017; Satterstrom et al., Cell, 2020. All papers are cited in text. Additionally, expression data used for this work is openly available and may be obtained from GTEx v8: https://gtexportal.org/home/

