## Supplementary Information for "De novo mutation hotspots in homologous protein domains identify function-altering mutations in neurodevelopmental disorders"

Supplementary information to “De novo mutation hotspots in homologous protein domains identify a distinct class of neurodevelopmental disorder genes and point to new candidate genes”

#### Supplementary Data

##### Supplementary Data S1. De novo mutations

Supp_data_S1.DDD_RUMC_GDX_denovos_filtered_31058_ntrios_fp_removed_2019_05_15

##### Supplementary Data S2. De novo mutation missense hotspot results

Supp_Data_S2_missense_DNM_hotspot_results.xlsx

##### Supplementary Data S3. De novo mutations at significant hotspot

Supp_data_S3_variants_at_significant_unique_count_hotspots_VEP_annotated.xlsx

##### Supplementary Data S4. All variants at hotspots

Supp_Data_S4_all_variants_from_samples_identified_by_restricted_count_method.xlsx

##### Supplementary Data S5. De novo mutation synonymous hotspot results

Supp_Data_S5_synonymous_DNM_hotspot_results.xlsx

##### Supplementary Data S6. De novo mutation nonsense hotspot results

Supp_Data_S6_nonsense_DNM_hotspot_results.xlsx

##### Supplementary Data S7. YASARA structures

Supp_data_S7_YASARA_structures_for_unique_count_hotspots.sce

**Note:** These structures cannot be made available through medRxiv. Please contact the corresponding author for this data.

##### Supplementary Data S8. Structural effects of missense DNMs

Supp_data_S8_hotspot_variants_structural_effects_annotated.xlsx

##### Supplementary Data S9. Mutational constraint in hotspot and proposed novel hotspot genes

Supplementary_Data_S9_Constraint.txt

##### Supplementary Data S10. Proportion of hotspot genes expressed across tissues

Supplementary_Data_S10_Proportion_hotspot_genes_expressed_across_tissues_all.txt

##### Supplementary Data S11. Proportion of hotspot genes expressed across tissues, PF00520 domain-containing genes

Supplementary_Data_S11_Proportion_hotspot_genes_expressed_across_tissues_PF00520.txt

##### Supplementary Data S12. Probability density functions for the classification of proposed novel hotspot genes

Supplementary_Data_S12_PDFs_for_proposed_novel_hotspot_classification.txt

##### Supplementary Data S13. Variation at stringent hotspot positions in clinical databases

Supplementary_Data_S12_Variation_at_hotspot_positions_ClinVar_HGMD_VGKL_Swissprot.txt

##### Supplementary Data S14. Phenotypes of patients with missense mutations at hotspot positions

Supplementary_Data_S13_Phenotypes_of_patients_with_hotspot_missense.xlsx

##### Supplementary Data S15. Gene sets used in analysis

Supplementary_Data_S14_Gene_sets_all.txt

##### Supplementary Data S16. PF00520 domain-containing genes used in analysis

Supplementary_Data_S15_Gene_sets_PF00520.txt

#### Supplementary Figures

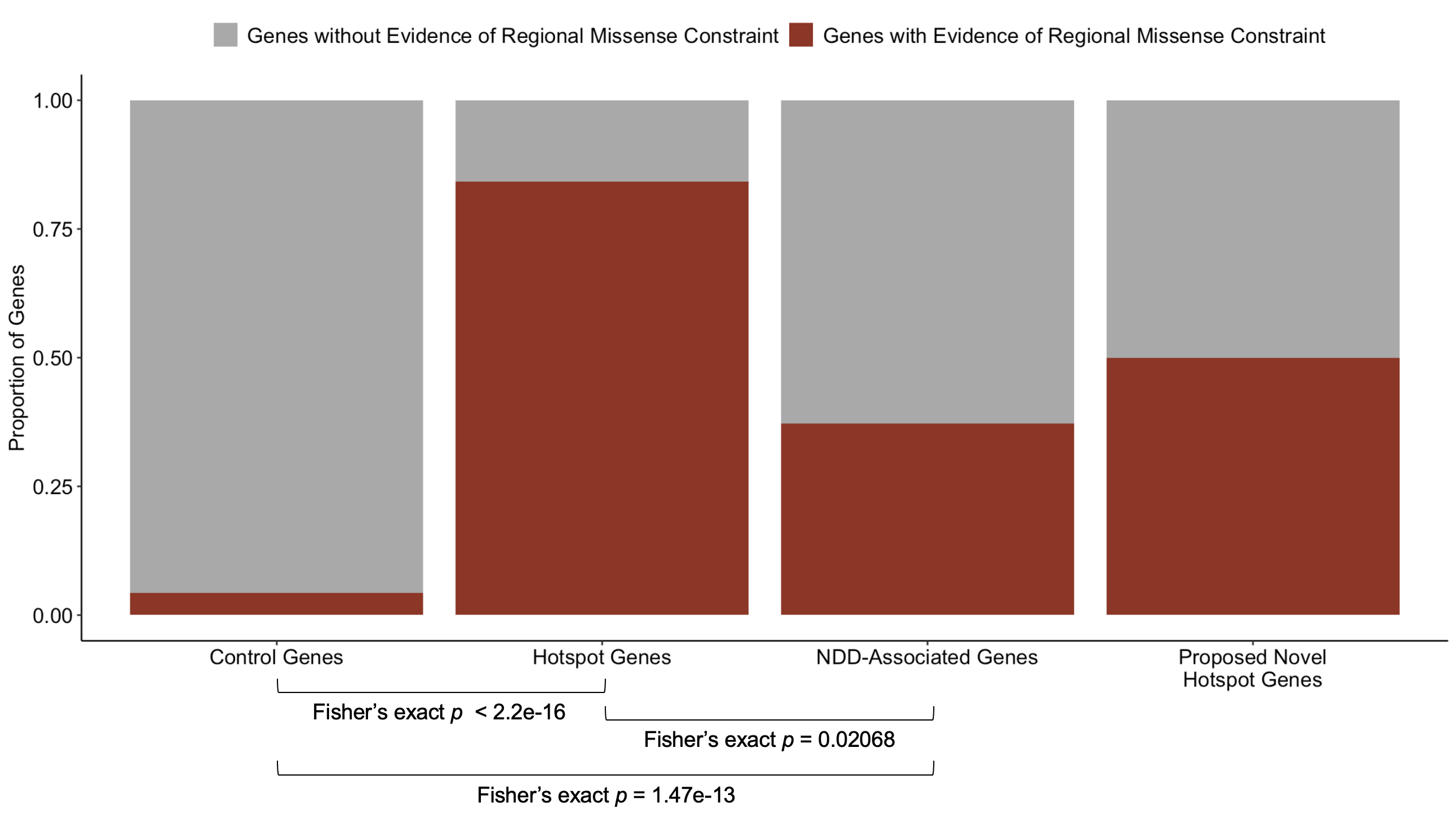

##### Supplementary Figure 1. A significant proportion of hotspot genes have evidence of regional missense constraint compared to control and NDD-associated genes.

Genes with evidence of regional missense constraint were taken from Samocha *et al.* (see **Methods**).^1^ The proportion of genes with and without evidence of regional missense constraint in this list were compared for control genes, NDD-associated genes, hotspot genes, and proposed novel hotspot genes. Hotspot genes have a significantly higher proportion of genes with regional missense constraint compared to control genes (Fisher’s exact p < 2.2 x 10^-16^) and other NDD-associated genes (Fisher’s exact p = 0.02).

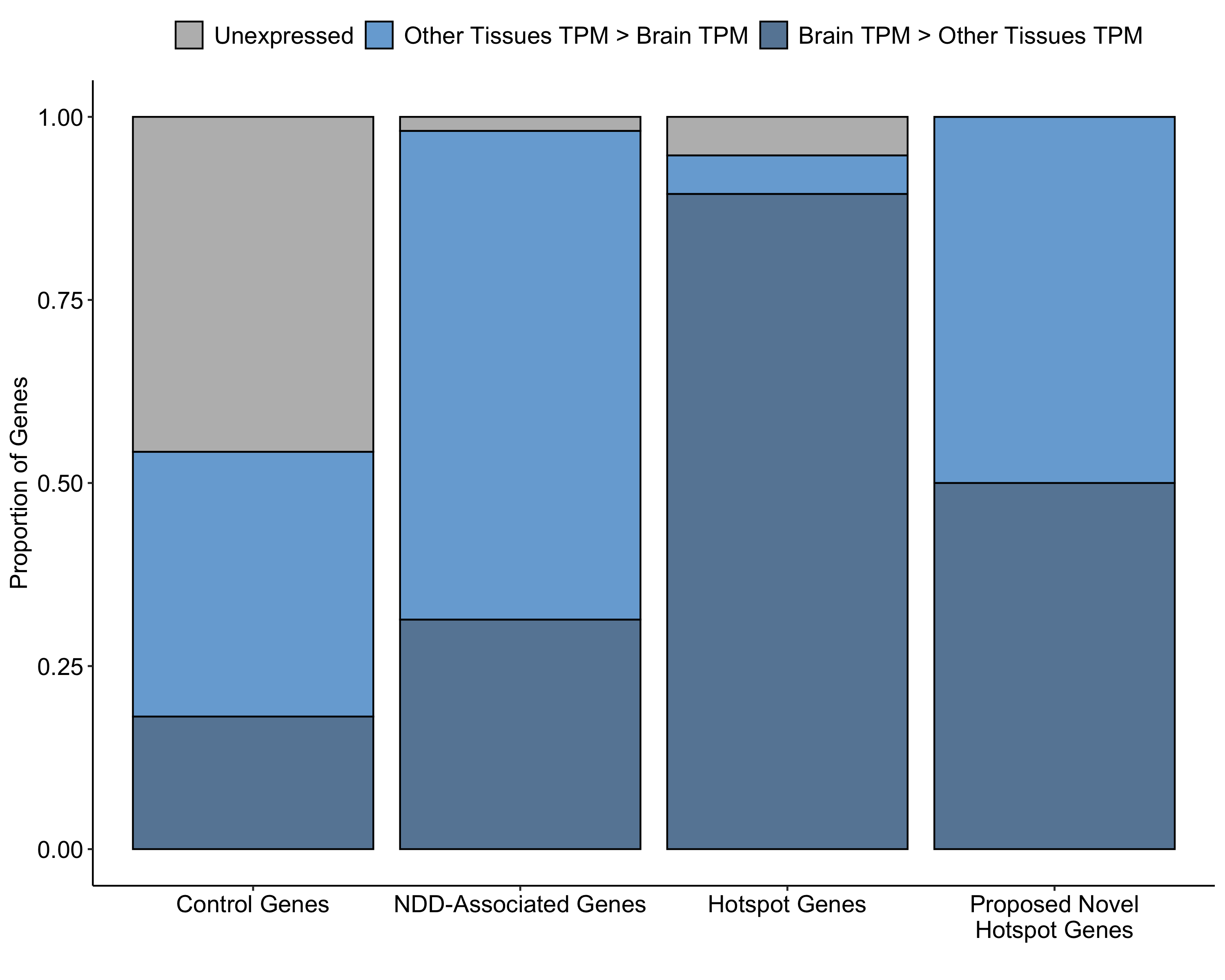

##### Supplementary Figure 2. A higher proportion of hotspot genes are expressed in brain than NDD-associated or control genes.

We compared the proportion of unexpressed genes (grey), genes expressed higher in other tissues than in brain by median TPM (light blue), and genes expressed higher in brain than in other tissues by median TPM (dark blue) across four gene sets (control genes, NDD-associated genes, hotspot genes, and proposed novel hotspot genes, see **Methods**). A significantly greater proportion of hotspot genes are expressed in brain than control genes (Fisher’s exact p = 2.985 x 10^-5^) and NDD-associated genes (Fisher’s exact p = 0.002).

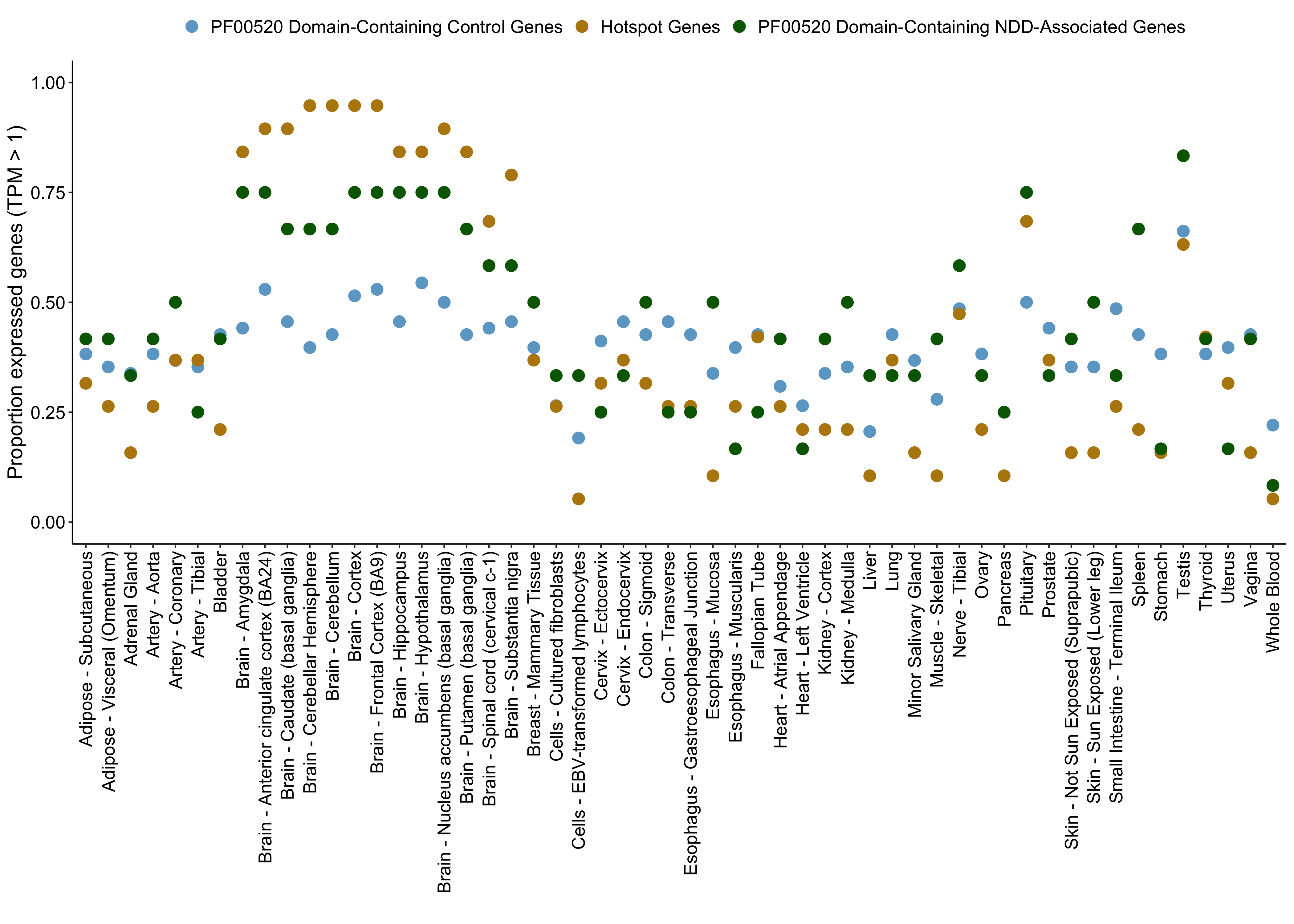

##### Supplementary Figure 3. Proportion of hotspot genes expressed across tissues compared to PF00520 domain-containing NDD-associated genes and PF00520 domain-containing control genes.

To determine whether the unique expression profile we observed for our hotspot genes was characteristic of all PF00520 domain-containing genes, we compared hotspot genes to NDD-associated genes containing a PF0050 domain (green, n = 12) and control genes containing a PF00520 domain (blue, n = 68) without sampling. A significantly greater proportion of hotspot genes are expressed in the caudate (basal ganglia), cerebellar hemisphere, cerebellum, cortex, and frontal cortex (BA9) compared to control genes (see **Supplementary Data S11** for Bonferroni-corrected Fisher’s exact p-values across all tissues). We find no significant differences between NDD-associated genes containing a PF00520 domain and hotspot genes (**Supplementary Data S11**). We conclude that most NDD-associated PF00520 domain containing genes (n = 31) are expressed in brain, and we have statistical power to detect mutation hotspots in 19 of these genes.

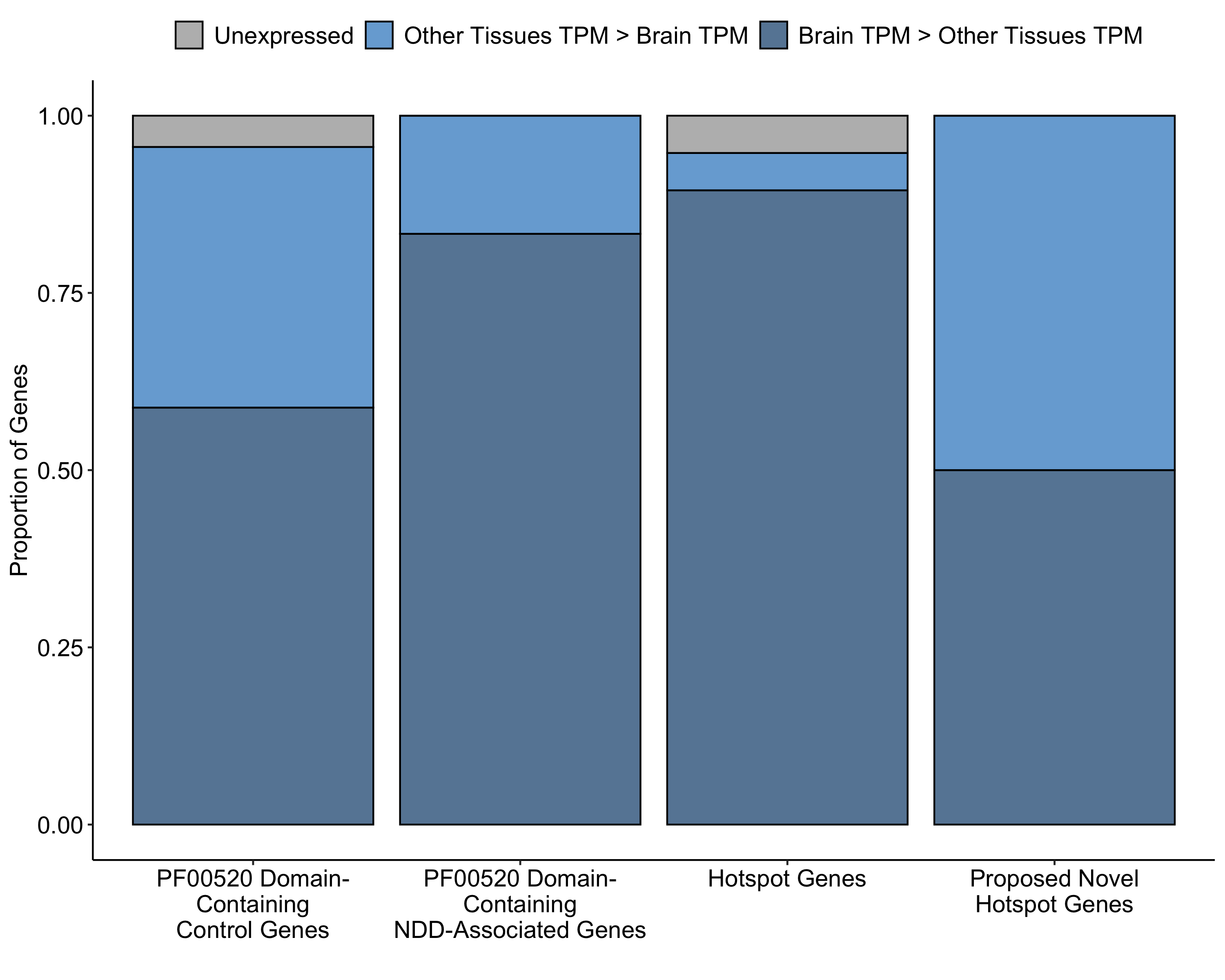

##### Supplementary Figure 4. A higher proportion of hotspot genes are expressed in brain than PF00520 domain-containing control genes.

In addition to looking at the proportion of genes expressed in a given tissue, we also considered whether hotspot genes were enriched for higher expression in brain than in other tissues. We show that a significant proportion of hotspot genes have higher expression in brain than in other tissues compared to control genes containing a PF00520 domain (Fisher’s exact p = 0.008), but not NDD-associated genes also containing this domain (Fisher’s exact p = 0.54). Hotspot genes likely represent a subset of NDD-associated PF00520 domain-containing genes, and all genes of this class could harbour pathogenic variation at hotspot positions.

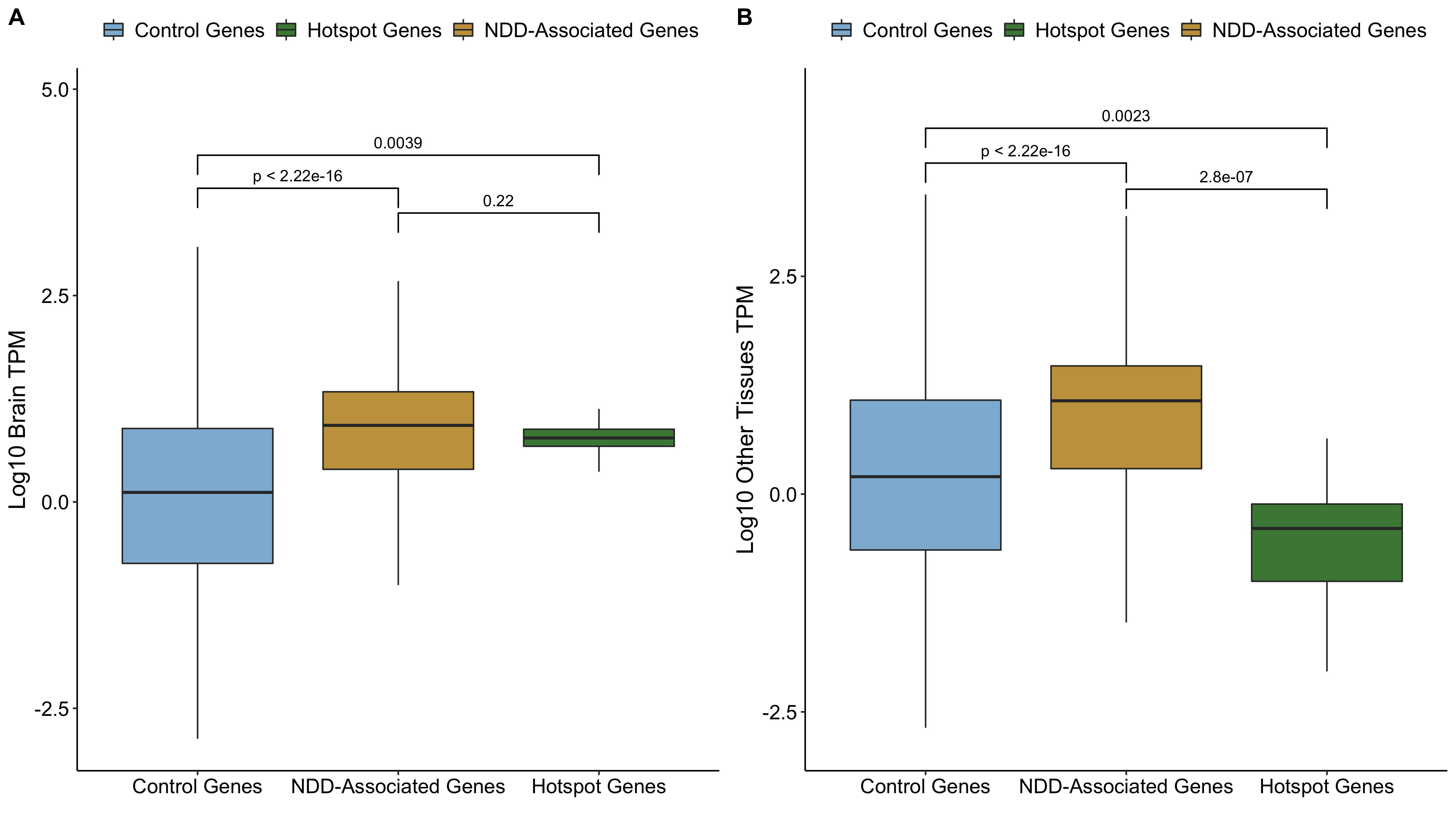

##### Supplementary Figure 5. TPM differences between hotspot, NDD-associated, and control genes in brain and other tissues.

We compared the median TPM distribution in brain (A) and other tissues (B) in expressed (TPM > 1) control, NDD-associated, and hotspot genes. We show that both NDD-associated and hotspot genes have higher expression in brain than control genes (Wilcoxon p < 2.2 x 10^-16^; Wilcoxon p = 0.0039). We also show that hotspot genes have significantly lower expression in other tissues compared to both control genes (Wilcoxon p = 0.0023) and NDD-associated genes (Wilcoxon p < 2.2 x 10^-16^). We use these expression differences to associate proposed novel hotspot genes with NDDs (see **Methods**).

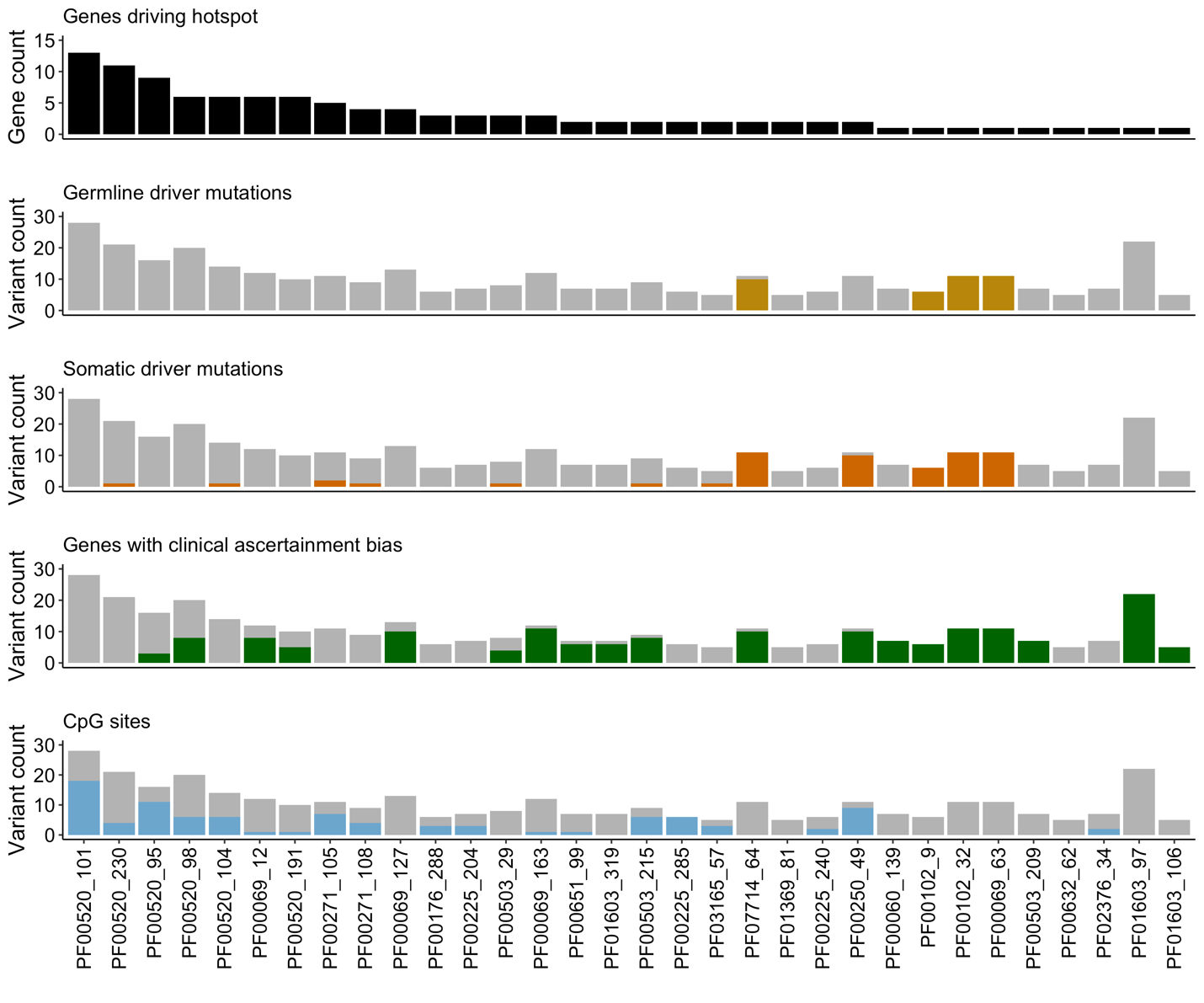

##### Supplementary Figure 6. Lenient hotspots may be driven by germline or somatic driver mutations, clinical ascertainment bias, and CpG hypermutability

Lenient hotspots may be driven by variants at the same protein consensus position but different genetic positions, the same genetic position recurrently mutated, or both. Kaplanis *et al.* describe recurrent missense variants as those mutated > 9 times in our cohort, and show that these are driven by four major processes: mutations that confer a proliferative advantage in the germline (germline drivers), mutations that confer a proliferative advantage in somatic tissues (somatic drivers), biases in clinical ascertainment and CpG hypermutability. We considered which of these factors might be driving our lenient mutation hotspots (sorted by the number of genes with mutations at the hotspot, black, top panel) by considering the proportion of mutations at each position driven by these four factors. Mutations in genes known to confer a proliferative advantage in the germline (second panel, yellow) and in the somatic tissue (third panel, orange) are coloured as a proportion of the total number of missense variants at the hotspot. Similarly, genes with clinical ascertainment bias – described here as those in the top 5% of the recurrent missense variant distribution – are coloured in green (fourth panel), and mutations at CpG sites are coloured blue (fifth panel).

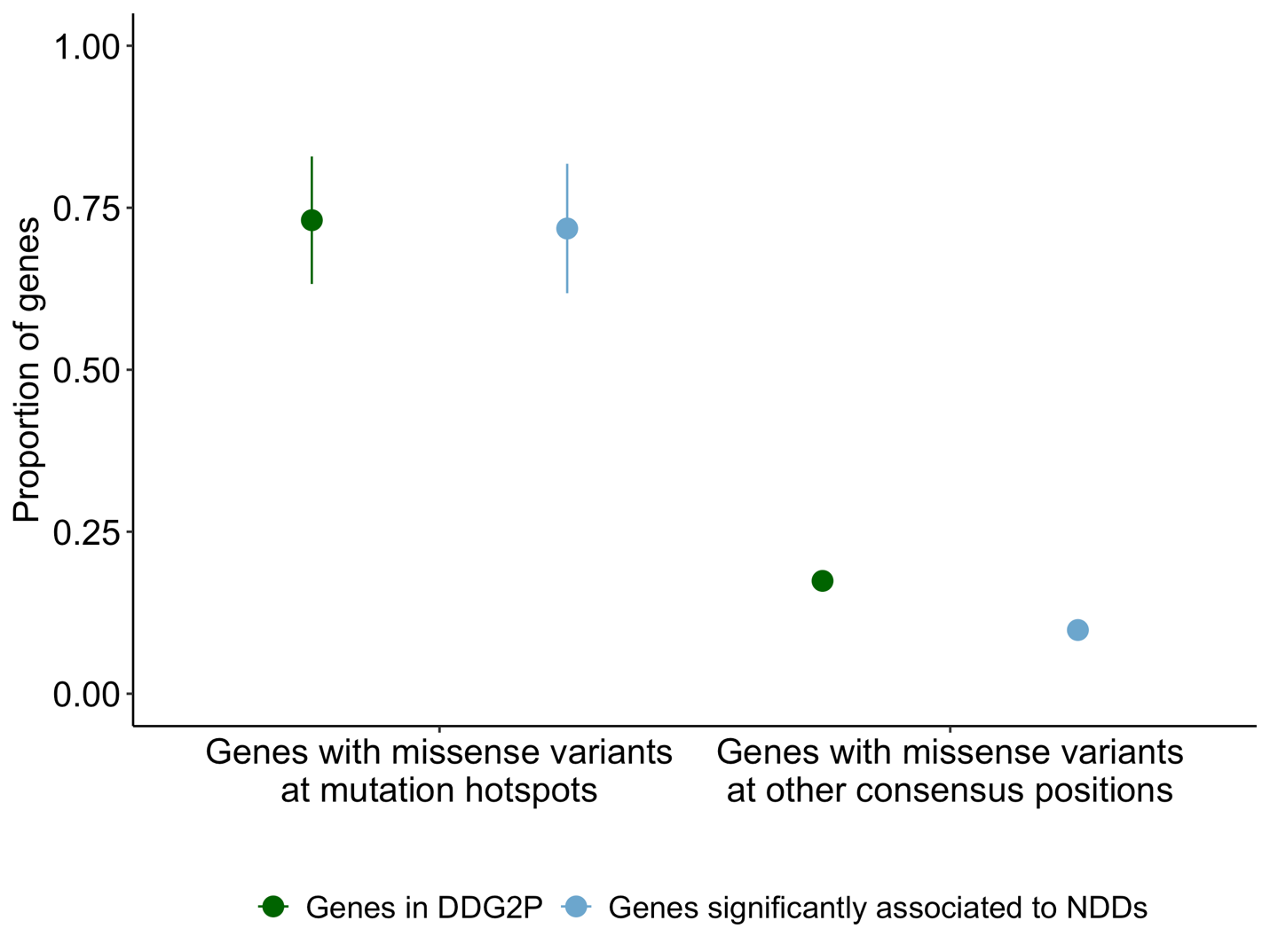

##### Supplementary Figure 7. Lenient hotspots are enriched for NDD-associated and DDG2P genes

The proportion of lenient hotspot missense variants in genes statistically associated to NDDs (blue) and in DDG2P (green) is shown at mutation hotspots (left) and all other protein consensus positions (right). Mutation hotspots are significantly enriched for missense mutations in genes statistically associated to NDDs (Fisher’s exact *p* < 2.2 x 10^-16^) and in DDG2P (Fisher’s exact *p* < 2.2 x 10^-16^).

#### Supplementary Tables

##### Supplementary Table 1 – NDD DNMs after processing

|  | **Original** | **MetaDomain Annotated** | **Located in Pfam Protein Domain** | **Meta-Domain Position Annotated** |
| --- | --- | --- | --- | --- |
| **Missense** | 28,241 | 26,178 | 13,114 | 11,288 |
| **Synonymous** | 9,005 | 8,496 | 3,862 | 3,229 |
| **Stop-gained** | 2,685 | 2,415 | 926 | 805 |
| **Total** | 39,931 | 37,089 | 17,902 | 15,322 |

Description of DNMs from Kaplanis et. al. study^4^ after DNM annotation and filtering (see **Methods**).

##### Supplementary Table 2 – Missense variant counts at hotspot positions p.96, p.102, p.231

| **Hotspot Position** | **Total Missense Variants at Position** | **Unique Missense Variants at Position** |
| --- | --- | --- |
| p.96 | 16 | 10 |
| p.102 | 20 | 13 |
| p.231 | 21 | 14 |

The number of missense variants at each hotspot position is summarised. The total missense variants represent all variants at the protein consensus position, including identical variants. Unique variants are counted as all unique chromosome, position, ref, alt at a protein consensus position without the inclusion of identical variants.

##### Supplementary Table 3 – Genes with missense DNMs hotspots by unique counting

|  | **With Missense DNMs at Significant Hotspot** | **Without Missense DNMs at Significant Hotspot** | **Total** |
| --- | --- | --- | --- |
| **DD-associated Genes** | 19 | 596 | 615 |
| **Other Genes** | 6 | 4,998 | 5,004 |
| **Total** | 25 | 5,594 | 5,619 |

A comparison of NDD-associated genes and genes not associated to NDD from the perspective of significant missense DNM identified via unique counting of DNMs. Contingency table (Chi-square p = *1.11^-13^*, test-statistic = 55.17, degrees of freedom = 1) featuring counts of genes that have missense DNMs in a potential hotspot location: i.e. located at a position that can be aggregated via homologous protein domain relations. Both the missense DNMs and diagnostic lists result from the Kaplanis et al. study.^4^ Based on this data, NDD-associated genes are by a 3.17 fold more likely to have a significant missense DNM hotspot than genes that do not have NDD-association.

##### Supplementary Table 4 – Hotspot genes are enriched for gain-of-function mutation consequences in DDG2P

|  | **Gain of Function Mutation Consequence** | **Other Mutation Consequence** |
| --- | --- | --- |
| **Hotspot genes in DDG2P** | 6 | 10 |
| **Other DDG2P Genes** | 163 | 1967 |

*Hotspot genes were tested for an enrichment of gain of function or activating mutation consequences (see* ***Methods****). Genes can belong to only one class (hotspot or other DDG2P genes), but their mutation consequences are considered independent (they can have both a gain of function mutation consequence and a different mutation consequence provided they are both in DDG2P). Gain of function mutation consequences were enriched in the hotspot gene set in DDG2P compared to other genes (Fisher’s exact p-value = 5.484 x 10^-5^).*

##### Supplementary Table 5 – NDD-associated genes have higher levels of constitutive expression than control genes

|  | **Constitutively Expressed** | **Not Constitutively Expressed** | **Unexpressed** | **Total Not Constitutively Expressed** |
| --- | --- | --- | --- | --- |
| **Control Genes** | 7853 | 23052 | 24278 | 47330 |
| **NDD-Associated Genes** | 476 | 505 | 11 | 516 |

*To show that NDD-associated genes generally have higher constitutive expression than control genes, we counted constitutively expressed (TPM > 1 in all tissues) and not constitutively expressed (TPM <= 1 in all tissues) genes in each set in GTEx data. NDD-associated genes have significantly higher levels of constitutive expression than control genes, even if we just consider genes in both sets that are expressed (TPM > 1 in at least one tissue; Fisher’s exact p < 2.2 x 10^-16^ in both sets).*

##### Supplementary Table 6 – Lenient hotspot positions are enriched for likely pathogenic missense variation in clinical databases

**VKGL:**

|  | **Hotspot consensus positions** | **Other consensus positions** |
| --- | --- | --- |
| **Likely pathogenic variants** | 61 | 3314 |
| **Likely benign variants** | 3 | 9465 |

Fisher’s exact *p* < 2.2 x 10^-16^

|  | **Hotspot consensus positions (no DNM at position)** | **Other consensus positions**  **(no DNM at position)** |
| --- | --- | --- |
| **Likely pathogenic variants** | 32 | 3154 |
| **Likely benign variants** | 3 | 9429 |

Fisher’s exact *p* < 2.2 x 10^-16^

|  | **Hotspot consensus positions**  **(no DNM at codon)** | **Hotspot consensus positions**  **(no DNM at codon)** |
| --- | --- | --- |
| **Likely pathogenic variants** | 26 | 3096 |
| **Likely benign variants** | 3 | 9398 |

Fisher’s exact *p* = 3.08 x 10^-13^

**ClinVar:**

|  | **Hotspot consensus positions** | **Other consensus positions** |
| --- | --- | --- |
| **Likely pathogenic variants** | 176 | 12985 |
| **Likely benign variants** | 9 | 12335 |

Fisher’s exact *p* < 2.2 x 10^-16^

|  | **Hotspot consensus positions (no DNM at position)** | **Other consensus positions**  **(no DNM at position)** |
| --- | --- | --- |
| **Likely pathogenic variants** | 121 | 12074 |
| **Likely benign variants** | 9 | 12294 |

Fisher’s exact *p* < 2.2 x 10^-16^

|  | **Hotspot consensus positions**  **(no DNM at codon)** | **Hotspot consensus positions**  **(no DNM at codon)** |
| --- | --- | --- |
| **Likely pathogenic variants** | 104 | 11861 |
| **Likely benign variants** | 9 | 12254 |

Fisher’s exact *p* < 2.2 x 10^-16^

*We compared the proportion of likely pathogenic missense variants at hotspot positions versus all other protein consensus positions in VKGL (top) and ClinVar (bottom). We compared all positions (first table), positions without a DNM at our cohort (second table), and positions without a DNM in the codon in our cohort (third table). Statistical significance was calculated using Fisher’s exact test.*

##### Supplementary Table 7 – Lenient hotspots are significantly enriched for missense variants in NDD and ASD probands

|  | **Hotspot consensus position missense DNMs** | **Other consensus position missense DNMs** |
| --- | --- | --- |
| **NDD probands** | 335 | 11294 |
| **Unaffected individuals** | 3 | 1383 |

Fisher’s exact *p* = 3.5 x 10^-13^

|  | **Hotspot consensus position missense DNMs** | **Other consensus position missense DNMs** |
| --- | --- | --- |
| **ASD probands** | 19 | 1868 |
| **Unaffected individuals** | 3 | 1383 |

Fisher’s exact *p =* 0.007

|  | **Hotspot consensus position missense DNMs** | **Other consensus position missense DNMs** |
| --- | --- | --- |
| **CHD probands** | 6 | 736 |
| **Unaffected individuals** | 3 | 1383 |

Fisher’s exact *p* = 0.07

*We compared the number of missense DNMs at hotspot positions and other protein consensus positions in cohorts of affected probands (NDD, ASD, and CHD) compared to a set of healthy population controls. NDD and ASD probands have a significant enrichment of missense DNMs in hotspot positions (Fisher’s exact test).*

##### Supplementary Table 8 – Lenient hotspots are not significantly enriched for synonymous variants

|  | **Hotspot consensus position synonymous DNMs** | **Other consensus position synonymous DNMs** |
| --- | --- | --- |
| **NDD probands** | 4 | 3229 |
| **Unaffected individuals** | 0 | 530 |

Fisher’s exact *p* = 1

|  | **Hotspot consensus position synonymous DNMs** | **Other consensus position synonymous DNMs** |
| --- | --- | --- |
| **ASD probands** | 2 | 717 |
| **Unaffected individuals** | 0 | 530 |

Fisher’s exact *p =* 0.51

|  | **Hotspot consensus position synonymous DNMs** | **Other consensus position synonymous DNMs** |
| --- | --- | --- |
| **CHD probands** | 0 | 236 |
| **Unaffected individuals** | 0 | 530 |

Fisher’s exact *p* = 1

*We compared the number of synonymous DNMs at hotspot positions and other protein consensus positions in cohorts of affected probands (NDD, ASD, and CHD) compared to a set of healthy population controls. No cohort has a significant enrichment of missense DNMs in hotspot positions (Fisher’s exact test).*

##### Supplementary Table 9 – ASD probands are significantly enriched for unique missense variants at lenient mutation hotspots

|  | **Hotspot consensus position unique missense DNMs** | **Other consensus position unique missense DNMs** |
| --- | --- | --- |
| **ASD probands** | 13 | 1821 |
| **Unaffected individuals** | 3 | 1371 |

Fisher’s exact *p =* 0.047

|  | **Hotspot consensus position unique missense DNMs** | **Other consensus position unique missense DNMs** |
| --- | --- | --- |
| **CHD probands** | 0 | 714 |
| **Unaffected individuals** | 3 | 1371 |

Fisher’s exact *p* = 1

*We compared the number of unique missense DNMs at hotspot positions and other protein consensus positions in cohorts of affected probands (ASD and CHD) compared to a set of healthy population controls. ASD probands have a significant enrichment of unique missense DNMs in hotspot positions (Fisher’s exact test). We defined ‘unique DNMs’ as those not recurrent in any of the three datasets.*

##### Supplementary Table 10 – ACMG classification of DNMs located at stringent hotspots in genes without association to NDDs

| **Variant** | **ACMG classification** | **Additional Notes** |
| --- | --- | --- |
| *Chr11(GRCh37): g.2432929C>G; ENST00000452833.1;*  *c.2558G>C; p.850R>Q; PF00520:p.102;*  *TRPM5* [*604600](https://www.omim.org/entry/604600); | *Likely Pathogenic (Class 4)* | PS2: De novo (both maternity and paternity confirmed) in a patient with the disease and no family history  PM1: Located in a mutational hot spot and/or critical and well-established functional domain (e.g., active site of an enzyme) without benign variation  PP3: Multiple lines of computational evidence support a deleterious effect on the gene or gene product (conservation, evolutionary, splicing impact, etc.)    HOWEVER:  BS1: Allele frequency is greater than expected for disorder |
| *Chr11(GRCh37):g.68848911C>A;*  *ENST00000294309.3;*  *c.1734C>A; p.545R>S; PF00520:p.96;*  *TPCN2* [*612163](https://www.omim.org/entry/612163) | *Likely Pathogenic (Class 4)* | PS2: De novo (both maternity and paternity confirmed) in a patient with the disease and no family history  PM1: Located in a mutational hot spot and/or critical and well-established functional domain (e.g., active site of an enzyme) without benign variation  PM2: Absent from controls (or at extremely low frequency if recessive) in Exome Sequencing Project, 1000 Genomes Project, or Exome Aggregation Consortium  PP3: Multiple lines of computational evidence support a deleterious effect on the gene or gene product (conservation, evolutionary, splicing impact, etc.) |
| *Chr12(GRCh37):g.113706596G>A;*  *ENST00000550785.1 c.963G>A; p.265R>Q; PF00520:p.96;*  *TPCN1* [*609666](https://www.omim.org/entry/609666) | *Likely Pathogenic (Class 4)* | PS2: De novo (both maternity and paternity confirmed) in a patient with the disease and no family history  PM1: Located in a mutational hot spot and/or critical and well-established functional domain (e.g., active site of an enzyme) without benign variation  PP3: Multiple lines of computational evidence support a deleterious effect on the gene or gene product (conservation, evolutionary, splicing impact, etc.)    HOWEVER: BS1: Allele frequency is greater than expected for disorder |
| *Chr14(GRCh37):g.63417240C>T;*  *ENST00000322893.7; c.1249G>A; p.327R>H; PF00520:p.102;*  *KCNH5* [*605716](https://www.omim.org/entry/605716) | *Pathogenic (Class 5)* | PS2 De novo (both maternity and paternity confirmed) in a patient with the disease and no family history  PM1: Located in a mutational hot spot and/or critical and well-established functional domain (e.g., active site of an enzyme) without benign variation  PM2: Absent from controls (or at extremely low frequency if recessive) in Exome Sequencing Project, 1000 Genomes Project, or Exome Aggregation Consortium  PP3: Multiple lines of computational evidence support a deleterious effect on the gene or gene product (conservation, evolutionary, splicing impact, etc.)  PP5: Reputable source recently reports variant as pathogenic, but the evidence is not available to the laboratory to perform an independent evaluation |
| *Chr20(GRCh37):g.49621072C>T;ENST00000371571.4; c.1332G>A; p.349R>H; PF00520:p.102;*  *KCNG1* [*603788](https://www.omim.org/entry/603788) | *Likely Pathogenic (Class 4)* | PS2: De novo (both maternity and paternity confirmed) in a patient with the disease and no family history  PM1: Located in a mutational hot spot and/or critical and well-established functional domain (e.g., active site of an enzyme) without benign variation  PM2: Absent from controls (or at extremely low frequency if recessive) in Exome Sequencing Project, 1000 Genomes Project, or Exome Aggregation Consortium  PP3: Multiple lines of computational evidence support a deleterious effect on the gene or gene product (conservation, evolutionary, splicing impact, etc.) |
| *Chr9(GRCh37):g.140878675G>A;ENST00000371372.1; c.1887G>A; p.581R>H; PF00520:p.102;*  *CACNA1B* [*601012](https://www.omim.org/entry/601012) | *Pathogenic (Class 5)* | PS2: De novo (both maternity and paternity confirmed) in a patient with the disease and no family history  PM1: Located in a mutational hot spot and/or critical and well-established functional domain (e.g., active site of an enzyme) without benign variation  PM2: Absent from controls (or at extremely low frequency if recessive) in Exome Sequencing Project, 1000 Genomes Project, or Exome Aggregation Consortium  PP2: Missense variant in a gene that has a low rate of benign missense variation and in which missense variants are a common mechanism of disease  PP3: Multiple lines of computational evidence support a deleterious effect on the gene or gene product (conservation, evolutionary, splicing impact, etc.)    HOWEVER: 1 occurrence in gnomAD |

Pathogenicity classifications of the variants found at the hotspots that are located in genes that are not in the consensus and discordant gene lists of Kaplanis et al.^4^ obtained through variant curation by a laboratory specialist. Abbreviations are according to ACGM^5^ guidelines: BS, benign strong; BP, benign supporting; FH, family history; LOF, loss-of-function; MAF, minor allele frequency; path., pathogenic; PM, pathogenic moderate; PP, pathogenic supporting; PS, pathogenic strong; PVS, pathogenic very strong.

##### Supplementary Table 11 – Genes with lenient missense hotspots

|  | **With missense DNMs at significant hotspot** | **Without missense DNMs at significant hotspot** | **Total** |
| --- | --- | --- | --- |
| **NDD-Associated Genes** | 48 | 567 | 615 |
| **Other Genes** | 19 | 4,985 | 5,004 |
| **Total** | 67 | 5,552 | 5,619 |

A comparison of NDD-associated genes and genes not associated to NDD from the perspective of significant missense DNM hotspots identified via lenient counting of DNMs. Contingency table (Chi-square p = *1.26^-31^*, test-statistic = 136.92, degrees of freedom = 1) featuring counts of genes that have missense DNMs in a potential hotspot location: i.e. located at a position that can be aggregated via homologous protein domain relations. Both the missense DNMs and diagnostic lists result from the Kaplanis et al. study.^4^ Based on this data, NDD-associated genes are by a 2.53 fold more likely to have a significant missense DNM hotspot than genes that do not have NDD-association.

##### Supplementary Table 12 – Counts of PTV, missense, and synonymous variants in protein domains in external de novo mutation datasets

|  | **SNV PTVs** | **Missense variants** | **Synonymous variants** | **Total variants** |
| --- | --- | --- | --- | --- |
| **ASD**  **(Satterstrom *et al.*)** | 128 | 1883 | 714 | 2725 |
| **CHD**  **(Jin *et al.*)** | 45 | 741 | 235 | 1021 |
| **Unaffected**  **(Jonsson *et al.,***  **Satterstrom *et al.*)** | 60 | 1377 | 524 | 1961 |

All DNMs from Satterstrom et al. (autism-spectrum disorders, ASD), Jin et al. (congenital heart defects, CHD) and unaffected individuals (Jonsson et al., Satterstrom et al. unaffected siblings) were mapped to metadomains for our hotspot analysis. The number of SNV PTVs (stop_gained), missense variants, and synonymous variants in protein domains are shown per cohort.

### URLs

YASARA: <http://www.yasara.org/>

CATH-Gene3D: [http://www.cathdb.info/](http://www.cathdb.info/version/latest/superfamily/1.20.120.350)

MetaDome web server: <https://stuart.radboudumc.nl/metadome/>

MetaDome GitHub repository: <https://github.com/cmbi/metadome>

RCSB PDB: <http://www.rcsb.org>
